# The impact of atypical intrahospital transfers on patient outcomes: a mixed methods study

**DOI:** 10.1101/2023.04.26.23289016

**Authors:** Estera Mendelsohn, Kate Honeyford, Andy Brittin, Luca Mercuri, Robert Edward Klaber, Paul Expert, Céire Costelloe

## Abstract

**Background:** The architectural design of hospitals worldwide is centred around individual departments, which require the movement of patients between wards. However, patients do not always take the simplest route from admission to discharge, but can experience convoluted movement patterns, particularly when bed availability is low. Few studies have explored the impact of these rarer, atypical trajectories.

**Methods:** Using a mixed-method explanatory sequential study design, we firstly used three continuous years of electronic health record data prior to the Covid-19 pandemic, from 55,152 patients admitted to a London hospital network to define the ward specialities by patient type using the Herfindahl-Hirschman index. We explored the impact of ‘regular transfers’ between pairs of wards with shared specialities, ‘atypical transfers’ between pairs of wards with no shared specialities and ‘site transfers’ between pairs of wards in different hospital site locations, on length of stay, 30-day readmission and mortality. Secondly, to understand the possible reasons behind atypical transfers we conducted three focus groups and three interviews with site nurse practitioners and bed managers within the same hospital network.

**Results:** We found that at least one atypical transfer was experienced by 12.9% of patients. Each atypical transfer is associated with a larger increase in length of stay, 2.84 days (95%CI: 2.56-3.12), compared to regular transfers, 1.92 days (95%CI: 1.82-2.03). No association was found between odds of mortality, or 30-day readmission and atypical transfers after adjusting for confounders. Atypical transfers appear to be driven by complex patient conditions, a lack of hospital capacity, the need to reach specific services and facilities, and more exceptionally, rare events such as major incidents.

**Conclusion:** Our work provides an important first step in identifying unusual patient movement and its impacts on key patient outcomes using a system-wide, data-driven approach. The broader impact of moving patients between hospital wards, and possible downstream effects should be considered in hospital policy and service planning.

## BACKGROUND

The management of patients from hospital entry to exit is a major challenge in healthcare, amid bed reductions and an aging population.[1] Secondary health systems are often structured as wards within departmental disciplines, departments within hospitals, and hospitals within multi-site organisational networks (or hospital ‘trusts’ in the UK). Patients must move through various locations as their needs evolve, making intrahospital transfers a daily practice in health systems worldwide. Many initiatives have attempted to optimise this process, otherwise known as ‘patient flow’,[2], [3] typically by predicting demand on the most commonly used patient pathways, and managing the points of ‘constraint’ (i.e. where demand overwhelms the capacity) such as the emergency department (ED).[4]–[7] Often trade-offs exist between admitting patients to the most appropriate ward and accommodating all patients during peaks in bed demands. Patients can therefore undertake convoluted movement patterns, particularly when bed availability is low. The impact of these rarer, *atypical* trajectories is unclear. Despite the activity around expediting patient flow, few studies have analysed hospital-wide patient movement with an etiological approach which questions whether potential associations exist between specific transfer patterns and clinical outcomes.[2], [8]

Several studies have examined the link between the number of intrahospital transfers undergone by patients and adverse outcomes.[9] Taking a whole-system view of the patient journey, they show that patients with more intrahospital movements have worse outcomes with respect to length of stay (LOS), falls, infection risk and carers’ perceptions of patient discharge readiness.[10]–[16] However, intrahospital transfers occur for a variety of reasons, (e.g. isolation due to infection, transfer to higher level-of-care, procedures, bed pressures and patient preference) and this approach may overlook their unequal impacts on outcomes. From a more targeted perspective, the outcomes of ‘outlying’, ‘bed-spaced’ or ‘boarding’ individuals, which have been placed on clinically inappropriate wards have been assessed.[17] The practice of outlying individuals into a ward outside of their home speciality, resulting in transfers between inlier and outlier wards, can be a strategic decision to reduce ED congestion, and is reported across health systems.[18]–[20] By definition such patients deviate from the regular trajectory for their speciality. Some evidence suggests this increases LOS, subsequent readmissions, and mortality.[17] However, outliers are usually defined in a binary sense by whether the individual has been placed on an inappropriate ward, without consideration to their movements up to and beyond the outlying ward. This may be a significant source of unobserved confounding between outlying status and LOS, with intrahospital transfers shown to double LOS in some populations.[11] A second limitation is a lack of specificity in the definition of an outlying patient, which can be unclear, or simplified to medical patients on a surgical ward.[17] Relying on predefined speciality definitions may overlook occurrences where patients are on a suboptimal ward within their overarching division,[21] or misclassify patients admitted to wards with multi-speciality staff. These factors may lead to inaccurate effect estimates.

Patient movement connects many areas of the hospital and may lead to unintended consequences, aligning with common characterisations of a complex system.[22] However, while complexity can increase with the quantity and uniqueness of relationships between components,[23] the literature taking a whole-system view of patient transfers has not distinguished between transfer type, while the targeted outlier literature does not usually consider the whole patient trajectory. A combination of these two approaches is needed, which demarcates these more complex patient transfers from regular transfers, while maintaining a view of the whole patient hospital journey. Guided by a data-driven definition of ward specialities using EHR data, this two-strand study firstly defines atypical transfers as movements between wards with no overlapping specialities and explores their association with key patient outcomes. Secondly, to understand the nature of this novel exposure more fully, we explore the causes of atypical transfers using in-depth qualitative interviews and focus groups with site nurse practitioners and bed managers.

### Objectives

The overarching aim of this study is to understand the impact atypical transfers on patient outcomes, and why these transfers occur. Fulfilling this aim therefore requires both quantitative and qualitative data sources and is well suited to a mixed-methods approach.[24] The specific objectives from the quantitative and qualitative strands of the study are:

#### Quantitative objectives

- To provide a systematic, data-driven definition of atypical transfers
- To explore the differential effects of atypical movement patterns on the patient outcomes of: LOS, 30-day readmission and mortality

#### Qualitative objectives

- To identify the possible causes of atypical transfers based on site nurse practitioners’ and bed managers’ perceptions

## METHODS

An explanatory sequential mixed-methods study design, with a quantitative focus, was conducted using routinely collected quantitative EHR record hospital data and qualitative semi-structured focus groups and interviews. The quantitative data were collected and analysed under a retrospective cohort study design, while exploratory thematic content analysis was used to describe the factors underlying the quantitative results. Quantitative and qualitative findings were therefore integrated to generate an in-depth understanding of the atypical transfers exposure. An overview of the study design is given in Figure 1.

**Figure 1:**
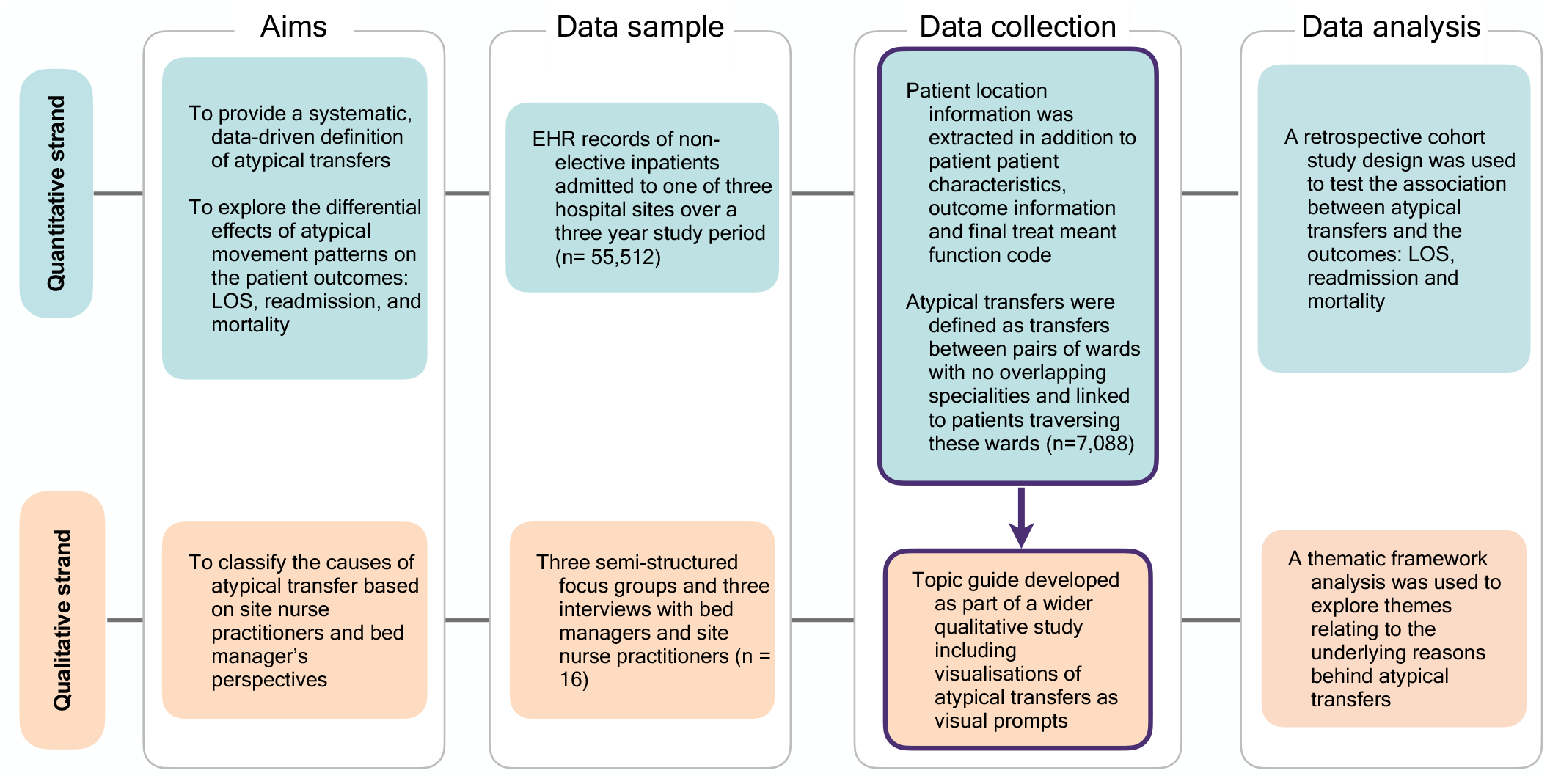
Overview of study design. An explanatory sequential study design was used, with the qualitative and quantitative strand interacting at the data collection stage. The qualitative findings were used to explain and enrich the quantitative findings.

### Retrospective Cohort Study

#### Study setting and participants

De-identified EHRs of patients admitted over a three-year period (falling between 2015 and 2018) were extracted. The data included patient and spell unique identifiers, while dates of admission were fully anonymised. The data structure and setting are described in detail elsewhere.[14] A retrospective cohort study design was used to examine the association between atypical ward transfers and the outcomes of LOS, 30-day readmission, and in-hospital mortality. Aiming to compare the relative effects of different types of patient movements, we excluded patients who had been treated on one ward for their entire spell, meaning the minimum exposure was therefore one intrahospital transfer. Our quantitative analysis therefore asks the question: among patients who move, does moving atypically increase length of stay, odds of readmission, or mortality? Maternity and paediatric patients were excluded (see Supplementary Note 1a). Likewise, elective patients were excluded as planned admissions exhibit a different acuity profile to emergency patients. The full inclusion criteria are shown in Supplementary Figure 1 and pre-analysis data processing is outlined in Supplementary Figure 2.

##### Exposure variables

Patient ward changes were defined as any change of location in the patients EHR including temporary movements to procedure wards but excluding informal movements the emergency department. The phrases patient transfer and movement are used interchangeably. Three types of patient transfers were considered: atypical, regular and site transfers (see Figure 2 for descriptions).

**Figure 2:**
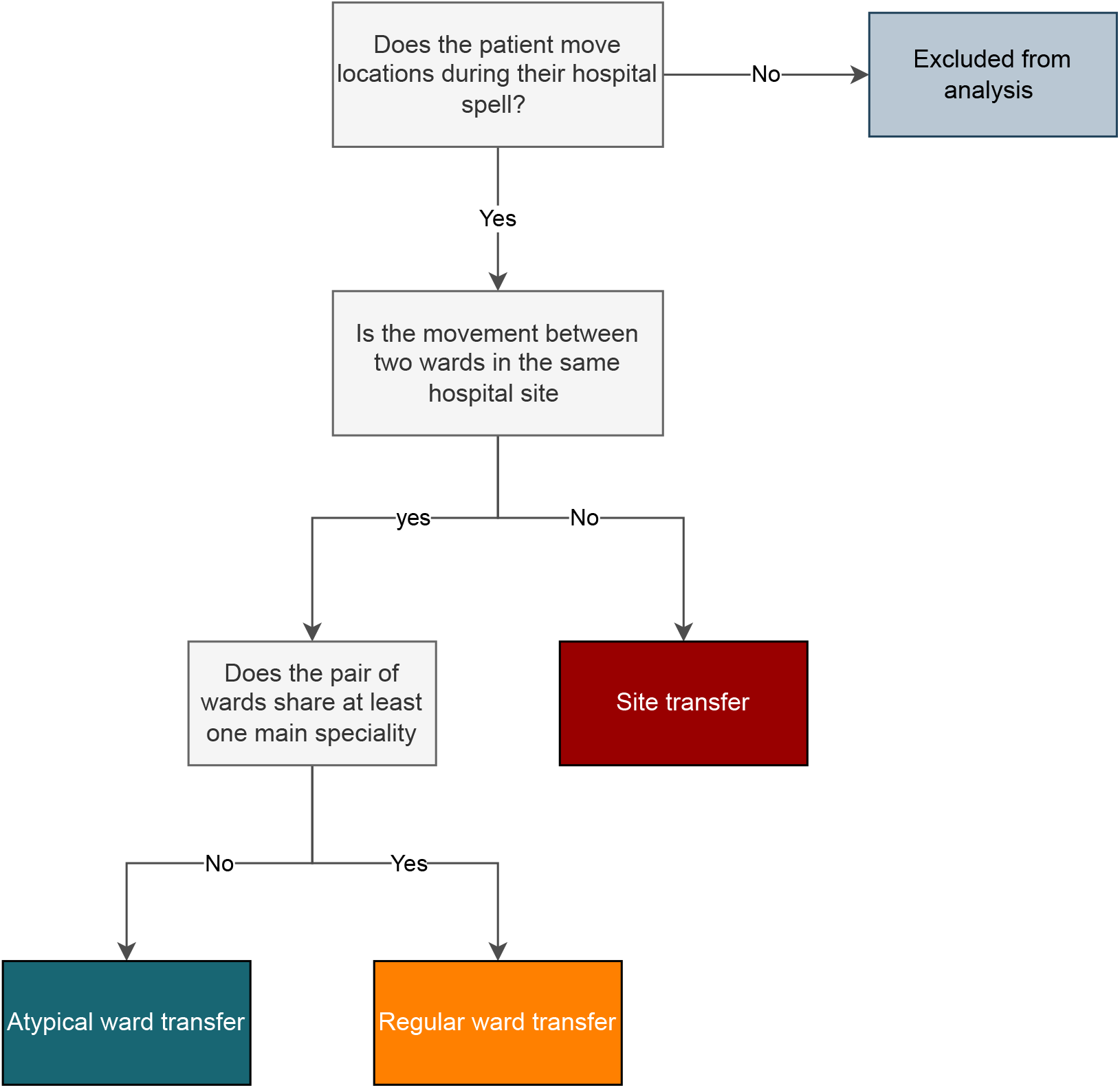
Decision tree flow chart outlining the criteria for the main exposures of interest: atypical transfers, regular transfers, and site transfers.

##### Ward specialities

We defined ward specialities based on the true hospital usage of the whole patient population, including elective and day-only patients. A concise description of the patient’s condition was first obtained using Treatment Function Codes (TFCs), which refer to the clinical division of the dominant healthcare professional responsible for the patient during an episode of care.[25] TFCs are time-dependent, with broader admitting specialities (e.g., General Medicine) typically evolving into more specific descriptions (e.g., Cardiology) as diagnoses and treatments are decided. The last recorded TFC was assumed to be the most accurate summary of the patient’s condition and selected for analysis. The patient’s final TFC was recorded against each unique ward entered during the spell, such that the patient ‘deposited’ their specialty at each stage of their journey through the hospital. Frequencies of TFCs were generated per ward, and a commonly used measure of market competitiveness, the Herfindahl-Hirschman Index (HHI),[26] was used to create a speciality diversity index, defined as:

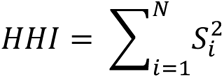

where *S*_i_ is the ‘share’ or proportion of patients admitted to a given ward from speciality *i*, and N is the total number of main specialities observed on the ward. The inverse of the HHI index is a measure of the ‘effective number of’ groupings, or the equivalent market size (EMS) corresponding to patient specialties ranked in descending order.[27] Each ward was assigned a number of representative specialties corresponding to their EMS rounded to the nearest integer. A high EMS indicates a multifunctional ward, while a low EMS indicates a highly specialist ward. Wards assigned the same specialties are not necessarily equivalent in function but could indicate a regular patient movement trajectory across two wards which deliver different services.

##### Atypical transfers

Atypical transfers were defined as a transfer between pairs of wards with no overlapping specialities identified from their EMS (see Figure 3). We therefore use the term atypical neutrally, reflecting the fact that such movements were uncommon given the speciality profiles of the two wards, rather than an appraisal on the appropriateness of the transfer at the individual patient level. To avoid inflating atypical transfer count by reciprocal trips to and from one ward, wards which only admitted patients for an average of 6 hours or less were verified and removed from the atypical transfer list if functioning as a temporary minor procedure ward, such as endoscopy. All other transfers were regarded as regular (non-atypical) transfers, with the exception site transfers (see Figure 2).

**Figure 3:**
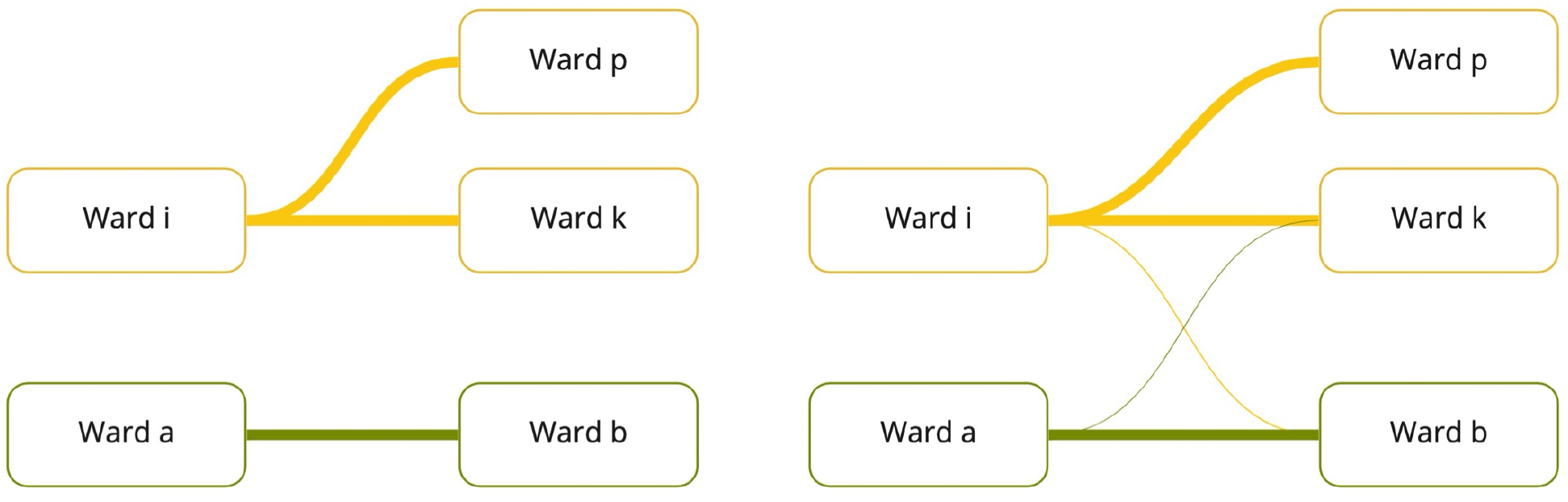
A depiction of a regular and atypical transfer movement. Ward pairs (*i,k),* and (*i,p)* belong to the same overarching speciality, making all transfers between them ‘regular’. Similarly, ward pairs (*a,b)* belong to an alternative overarching speciality and transfers between this pair are ‘regular’. Meanwhile, transfers between crossing specialities (such as (*a,k),* and (*i,b)*) are classed as ‘atypical’.

##### Outcome variables

Three outcomes were investigated: LOS, 30-day readmission and in-hospital mortality. A continuous variable for LOS was derived for each spell using the difference between admission and discharge timestamps (recorded up to 15 minutes resolution) converted to days and fraction of days. Unplanned readmissions were defined as emergency readmissions for any reason (except for pregnancy) within 30 days of an index hospitalisation. Index admissions were defined iteratively, such that each spell could became an index admission if followed by a subsequent admission. The unit of analysis in this regression was the index admission, and all covariates are taken from this spell. As all hospital sites pertained to the same hospital network, readmissions to different sites were included in the analysis (and distinguished from site transfers which were categorised under the same spell ID).[28] If the index hospitalisation ended in death, it was excluded from the analysis of readmission. Finally, in-hospital mortality was recorded as a binary variable if the spell ended in the patient’s death. The unit of analysis for all outcomes were hospital spells, therefore patients could contribute more than one hospitalisation to the main model if it fulfilled the selection criteria.

##### Confounding variables

We aimed to control for available covariates which confounded the exposure-outcome relationship.[29] Separate models were fitted for each outcome, and directed acyclic graphs, were used to guide variable selection (Supplementary Figure 3). As acutely unwell patients have been shown to have a higher number of transfers, and non-standard needs may be more prone to taking uncommon routes through the hospital,[30] patient acuity and complexity were considered confounders. We controlled for patient demographics and the following variables in the modelling: Elixhauser comorbidity index, admission to an ICU/critical care, weekend admissions and out of hours admissions (7pm-7am), the number of primary ICD-10 codes (as an estimate of multi-morbidity.[31]) the major diagnostic category of the first primary ICD-10 code recorded, discharge destination and severity of surgical procedures (diagnostic, minor, intermediate, and major procedures using existing code lists created by Abbott *et al.,* (see supplementary Note 1b) in addition to diagnostic imaging.[32])

#### Statistical Analysis

Atypical transfers were explored using network analysis and depicted using a chord diagram. Univariable regressions (see Supplementary Table 1a-c) and multivariable regression models were fitted separately. The association between atypical transfers and LOS was modelled by a generalized linear model (GLM) using a gamma distribution and a log-link. Average marginal effects (AMEs) were computed with respect to the variables of interest, holding all other variables constant.[33] It can be interpreted as the impact of a change in a focal independent variable on predicted value of the outcome, holding other variables constant.[34] AMEs were computed in R using the ‘margins’ command, specifying ‘type = response’.[35] An interaction term between atypical transfers and age was explored and found to be statistically insignificant. Logistic regressions were used to examine associations for the outcomes of mortality and 30-day readmission. Clustered standard errors by individual patient were implemented in all regression models using the Sandwich package. No major collinearity existed between variables. Large LOS ‘outliers’ were not removed, on the basis that these are true values in the data.[36], [37] Spells containing incomplete information were removed as missingness was minimal. The DHARMa package in R (Hartig 2018) was used to evaluate all model’s fit.

#### Sensitivity Analyses

We performed 6 sensitivity analyses to examine the robustness of the results by altering parameters related to the patient population, covariates exposure and model diagnostics (Table 1).

**Table 1:** Summary of sensitivity analyses:

| No. | Parameter | Modification |
| --- | --- | --- |
| 1 | **Patient population:**<br>discharged alive | We repeated the analysis of the impact of LOS with the exclusion of patients who did not survive their spell |
| 2 | **Patient population:**<br>negative for infection or colonisation | We omitted patients who tested positive for any type of infection, as this can significantly increase LOS, risk of death and readmission, and result in atypical transfers specifically due to isolation procedures |
| 3 | **Covariates:**<br>including approximate month of admission | The dates of admission anonymised and relative to a time 0, therefore seasonality effects could not be accurately investigated. Instead, an exploratory analysis was conducted to account for a possible seasonality effect via adjusting for a dummy variable created by binning every 30 days into 12 equal categories iterating over the three-year dataset |
| 4 | **Covariates:**<br>including alternative surgical category | As no official categorisation of OPCS-4 surgical codes exists, a second analysis was conducted an alternative definition based on Bupa reimbursement code lists (see supplementary Note 1c for details) |
| 5 | **Exposure definition:** using first patient episode TFC | To understand the sensitivity of the atypical transfers definition, analyses were also repeated using the first TFC of the patient's admission. |
| 6 | **Model diagnostics:** excluding potentially influential outliers | We reported the main analyses with the exclusion of observations whose standardised residuals were above 3.[38] |

Analyses were conducted using R version 4.2.1

### Qualitative focus groups and interviews

#### Study population

A purposeful sampling strategy was adopted whereby the clinical study lead sent an invitation email to site nurse practitioners (clinical staff, denoted by prefix ‘S’ in participant quotations) and bed managers (non-clinical staff, denoted by prefix ‘B’ in participant quotations) of varying seniority at the study setting. Participants were prioritised by years of experience, due to the quantitative data being historic (collected between 2015-2019) and by familiarity with all three hospital sites. Three focus groups (consisting of 4 to 5 participants) and three one-to-one interviews were conducted (totalling 16 participants). On average, participants had 13.75 years of experience at the hospital trust. The number of focus groups chosen was a pragmatic decision based on the availability of participants in this role during the study period.

#### Data collection

All focus groups and interviews were conducted remotely via Microsoft Teams between 9 July 2022 and 29 October 2022. The focus groups duration was around 1 hour and interviews 40 minutes. The decision to use online data collection methods was due to the risk posed by the potential of Covid-19 transmission between participants, and the fact it allowed participants to join from different sites while minimising disruption to their working hours. A focus group topic guide which comprised visualisations of atypical transfer pathways was pilot tested and revised.

#### Data analysis

Interviews and focus groups were visually recorded, transcribed and checked for accuracy by EM. The thematic framework method was used to analyse the qualitative data content surrounding atypical transfers. Codes were inductively generated from the data, with the resulting codes used as the basis of a thematic framework. Subsequently, the framework was used to index, chart, map and interpret the data within and between cases.[39] A descriptive approach was taken, with themes remaining close to the participant’s accounts, and attention was paid to deviant cases which were included in the findings.[40] Annotation and coding of transcripts were conducted using NVivo (Version 12, QSR International, Burlington, Massachusetts, USA).

#### Ethical approval

This study was defined as service evaluation by the Health Service Research Authority and therefore NHS Research Ethics Committee approval was not needed. The study was approved as a service evaluation through Imperial College Healthcare NHS Trust (Ref:347/Ref:719) Ethical research practice standards were followed throughout, including obtaining informed consent and right to withdraw from the study at any point.

## RESULTS

### Patient characteristics

A total of 55,152 non-elective spells taking place during the 3-year study period met the entry criteria for the study, of which 7,088 (12.9%) experienced at least one atypical transfer between pairs of wards with no common main specialities (Figure 3). Of these, 5,844 (82.4%) undertook one atypical transfer, while 1,244 (17.6%) undertook two or more atypical transfers. Meanwhile, 8.5% of all patients experienced at least one site transfer. Over half of the population (54.2%) were transferred once during their spell (n=29,868). A breakdown of transfer type for patients with multiple transfers is given in Supplementary Figure 4. Most of the study population was male (52.9%), over 65-years old (55.2%), and 69.8% exhibited at least one Elixhauser comorbidity. Out-of-hours admissions were common, with 48.0% of admissions occurring between 7pm and 7am, while 23.8% of admissions occurred at weekends. The median LOS was 7.2 days, and 3,022 (5.5%) of spells resulted in in-hospital death.

Patients undertaking atypical transfers did not differ from those with no atypical transfers with respect to age, site transfers, Elixhauser comorbidities, or in-hospital death, with no statistically significant differences between non-atypical and atypical transfer patients (Table 2). However, atypical transfer patients were more likely to be admitted at weekends and out of hours (25.3% vs 23.5% p<0.001, and 52.0% vs 47.4% p<0.001, respectively). Such patients also underwent more procedures of all categories (excluding major procedures), but had fewer admissions to the intensive care unit (ICU) (5.8% vs 6.9%, p<0.001) (Table 2). A full descriptive characteristics table is given in Supplementary Table 2.

**Table 2:** Descriptive characteristics of the patient population, stratified by whether they undertook at least one atypical transfer or not.

|  | No atypical transfers<br>(n=48,064) | At least one atypical transfer<br>(n=7,088) | Total<br>(n=55,152) | p-value |
| --- | --- | --- | --- | --- |
| Regular ward transfers |  |  |  | < 0.001 |
| **Median** | 1 | 0 | 1 |  |
| **Q1, Q3** | 1, 2 | 0, 2 | 1, 2 |  |
| Site transfers |  |  |  | 0.677 |
| **Median** | 0 | 0 | 0 |  |
| **Q1, Q3** | 0, 0 | 0, 0 | 0, 0 |  |
| Age (years) |  |  |  | 0.245 |
| **18-39** | 6273 (13.1%) | 906 (12.8%) | 7179 (13.0%) |  |
| **40-65** | 15332 (31.9%) | 2205 (31.1%) | 17537 (31.8%) |  |
| **over 65** | 26459 (55.0%) | 3977 (56.1%) | 30436 (55.2%) |  |
| Gender |  |  |  | < 0.001 |
| **Female** | 22419 (46.6%) | 3589 (50.6%) | 26008 (47.2%) |  |
| **Male** | 25645 (53.4%) | 3499 (49.4%) | 29144 (52.8%) |  |
| Length of stay (days) |  |  |  | < 0.001 |
| **Median** | 7.0 | 9.0 | 7.2 |  |
| **Q1, Q3** | 3.3, 14.4 | 4.2, 19.6 | 3.5, 14.9 |  |
| Elixhauser comorbidities<br>(n, %) |  |  |  | 0.074 |
| **0** | 5254 (10.9%) | 839 (11.8%) | 6093 (11.0%) |  |
| **1-4** | 33591 (69.9%) | 4911 (69.3%) | 38502 (69.8%) |  |
| **>=5** | 9219 (19.2%) | 1338 (18.9%) | 10557 (19.1%) |  |
| Discharge destination |  |  |  | < 0.001 |
| **Not usual place of residence** | 7190 (15.0%) | 1212 (17.1%) | 8402 (15.2%) |  |
| **Usual place of residence** | 40874 (85.0%) | 5876 (82.9%) | 46750 (84.8%) |  |
| Primary diagnosis count<br>(n, %) |  |  |  | < 0.001 |
| **Mean (SD)** | 1.2 (0.4) | 1.3 (0.5) | 1.2 (0.4) |  |
| **Range** | 1.0 – 9.0 | 1.0 – 8.0 | 1.0 – 9.0 |  |
| Intensive Care Admission |  |  |  | < 0.001 |
| **No** | 44747 (93.1%) | 6678 (94.2%) | 51425 (93.2%) |  |
| **Yes** | 3317 (6.9%) | 410 (5.8%) | 3727 (6.8%) |  |
| In-hospital mortality |  |  |  | 0.187 |
| **No** | 45454 (94.6%) | 6676 (94.2%) | 52130 (94.5%) |  |
| **Yes** | 2610 (5.4%) | 412 (5.8%) | 3022 (5.5%) |  |
| Weekend admission |  |  |  | 0.001 |
| **No** | 36749 (76.5%) | 5294 (74.7%) | 42043 (76.2%) |  |
| **Yes** | 11315 (23.5%) | 1794 (25.3%) | 13109 (23.8%) |  |
| Out of hours admission (7pm-7am) |  |  |  | < 0.001 |
| **No** | 25273 (52.6%) | 3400 (48.0%) | 28673 (52.0%) |  |
| **Yes** | 22791 (47.4%) | 3688 (52.0%) | 26479 (48.0%) |  |
| Diagnostic imaging procedures (n, %) |  |  |  | < 0.001 |
| **0** | 23701 (49.3%) | 2646 (37.3%) | 26347 (47.8%) |  |
| **1-3** | 22206 (46.2%) | 3891 (54.9%) | 26097 (47.3%) |  |
| **4-6** | 1835 (3.8%) | 440 (6.2%) | 2275 (4.1%) |  |
| **Over 6** | 322 (0.7%) | 111 (1.6%) | 433 (0.8%) |  |
| Minor surgical procedures (n, %) |  |  |  | 0.008 |
| **0** | 40339 (83.9%) | 5881 (83.0%) | 46220 (83.8%) |  |
| **1-2** | 7193 (15.0%) | 1103 (15.6%) | 8296 (15.0%) |  |
| **3-5** | 473 (1.0%) | 87 (1.2%) | 560 (1.0%) |  |
| **Over 5** | 59 (0.1%) | 17 (0.2%) | 76 (0.1%) |  |
| Major surgical procedures (n, %) |  |  |  | < 0.001 |
| **0** | 41334 (86.0%) | 5857 (82.6%) | 47191 (85.6%) |  |
| **1-3** | 6198 (12.9%) | 1168 (16.5%) | 7366 (13.4%) |  |
| **4-6** | 453 (0.9%) | 55 (0.8%) | 508 (0.9%) |  |
| **Over 6** | 79 (0.2%) | 8 (0.1%) | 87 (0.2%) |  |
| Continuous variables were compared using the Kruskal–Wallis test, while categorical variables were compared by the chi-square test |  |  |  |  |

### Atypical transfer characteristics

Across the three hospital sites, 805 unique atypical transfers ward pairs were identified. While a small number of ward combinations account for the majority of atypical transfers, many atypical transfer routes rarely used, with 78% of pairs being used 10 or less times across the study period, and 29% of pairs only occurring once. Across all hospital sites the most travelled atypical paths included short-term observation wards such as assessment and clinical decision units (CDUs). While no transfers from A&E were recorded in the data, CDUs which are short stay wards under the care of emergency medicine consultants,[41] were labelled under the speciality A&E. This reflects the fact that most patients on this ward were discharged before seeing another consultant. Only 1.6% of all transfers involved the ICU or critical care ward, meaning that few atypical pathways are used to transport acutely deteriorating patients

### Multivariable regression results

After adjusting for the listed confounders and averaging over all observations in the data sample, each additional atypical transfer results in an estimated increase in LOS of 2.84 days (95% CI: 2.56-3.12). By comparison, regular ward transfers had an effect size of 1.92 days (95% CI: 1.82-2.03) increase in LOS for each additional transfer. Meanwhile, site transfers showed the largest effect on LOS with an increase of 3.02 days in LOS (95% CI:2.70-3.35) for each additional site transfer. Figure 5a summarises the AMEs from the multivariable GLM used for the focal predictors LOS.

**Figure 4:**
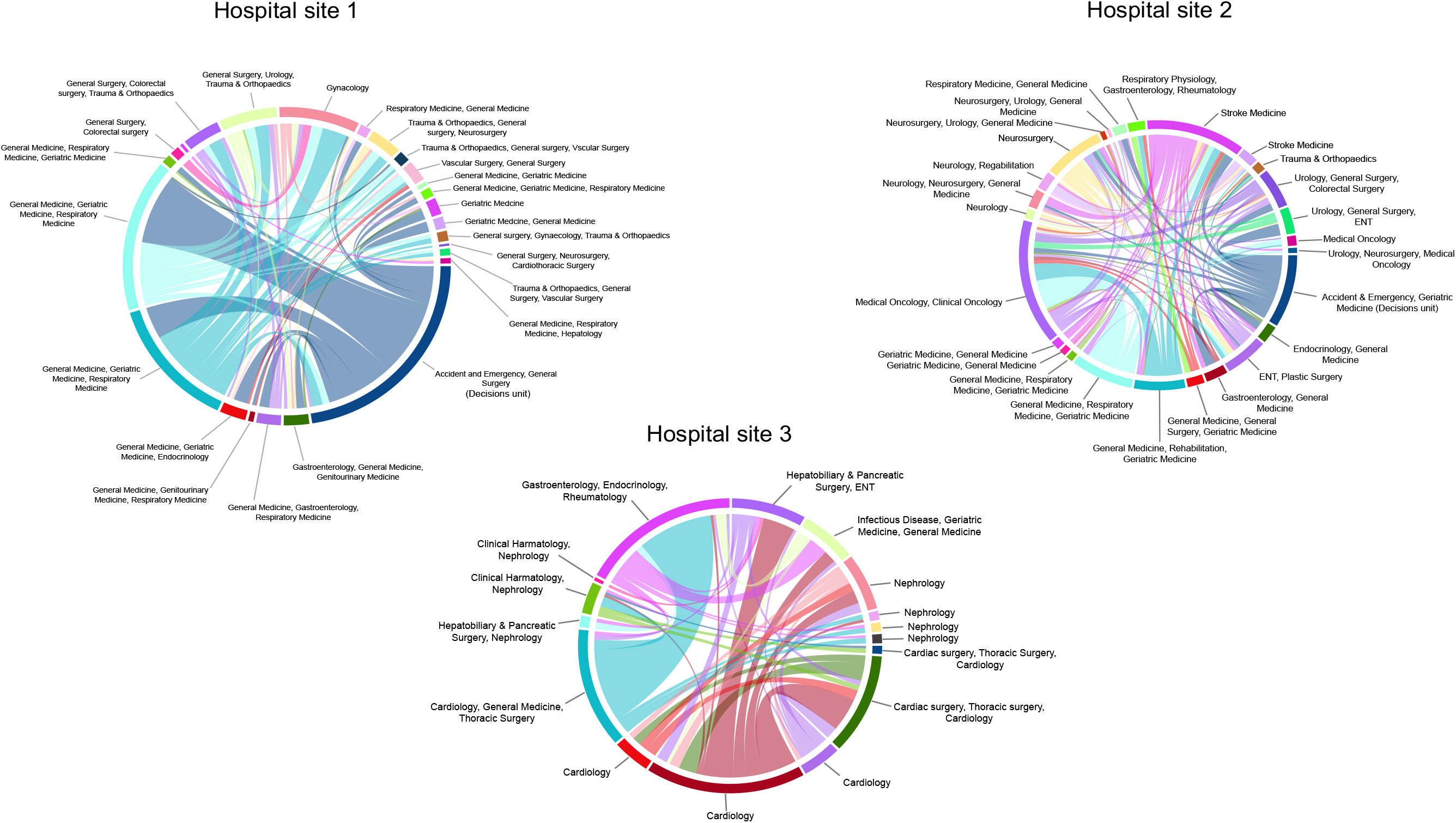
Chord diagrams depicting atypical transfers over the three hospital sites (A-C). Each sector in the outer track represents a ward, while each link between wards represents an atypical transfer. The thickness of the link is proportional to the volume of patients exchanged, between approximately 10 and 360 patients. The label of the track reflects the top specialities of the patients residing on that ward based on the whole patient population, with a maximum of 3 specialities.

**Figure 5:**
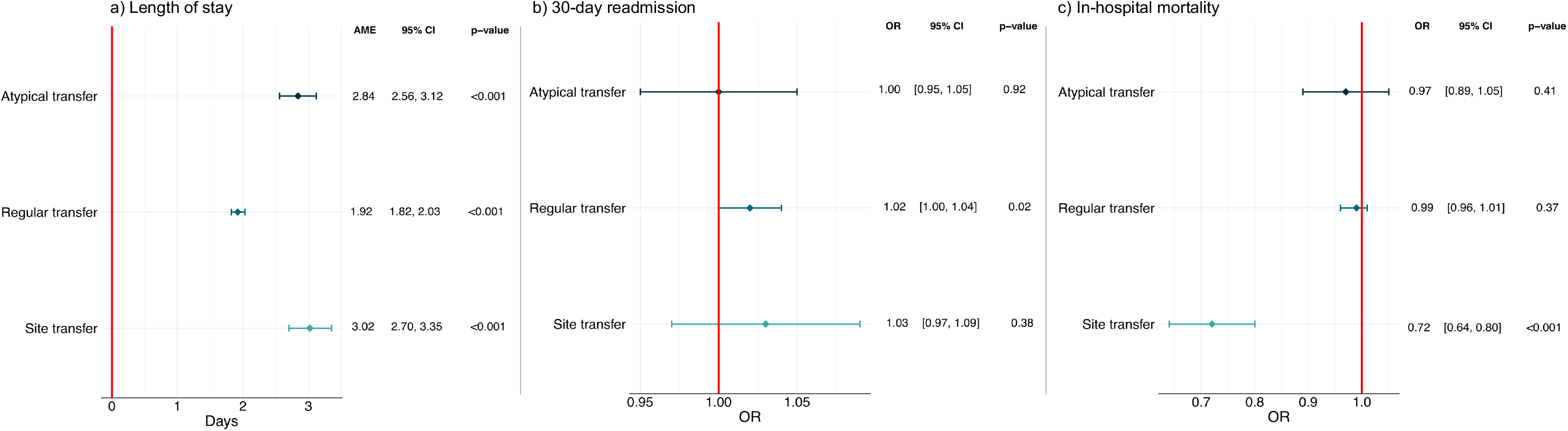
Multivariable regression results. Panel A shows the multivariable GLM regression with a gamma distribution and log-link predicting hospital LOS (n = 55,152). Results are presented as adjusted average marginal effects in days. In addition to atypical transfers, regular transfers, site transfers, the regression is controlled for age, gender, ethnicity, Elixhauser comorbidities, weekend admission, out of hours admission, discharge destination, admission to an ICU/critical care ward, number of primary diagnoses, imaging procedures, minor, intermediate and major surgical procedures, and major diagnostic category. Panel B shows a multivariable logistic regression predicting emergency readmission (n = 52,125), with results presented as adjusted odds ratios. All spells ending in in-hospital death were removed from this regression (n = 3,022) and 5 remaining patients were removed from the major diagnostic category ‘External causes of morbidity’ to avoid violating the positivity assumption.[39] In addition to atypical transfers, regular transfers, site transfers, the regression is controlled for age, gender, ethnicity, Elixhauser comorbidities, weekend admission, out of hours admission, discharge destination, admission to an ICU/critical care ward, number of primary diagnoses, imaging procedures, minor, intermediate and major surgical procedures, major diagnostic category and length of stay. Panel C shows a multivariable logistic regression predicting in-hospital mortality (n = 55,000), with results presented as adjusted odds ratios. Patients under the major diagnostic categories ‘external causes of morbidity’ and ‘factors influencing health status and contact with health services’ (n = 152) were similarly removed due to the positivity assumption. Covariates included in the model were: atypical transfers, regular transfers, site transfers, the regression is controlled for age, gender, ethnicity, Elixhauser comorbidities, weekend admission, out of hours admission, admission to an ICU/critical care ward, number of primary diagnoses, imaging procedures, minor, intermediate and major surgical procedures, and major diagnostic category. All 95% confidence intervals are based on standard errors clustered at the patient level.

Atypical transfers and site transfers did not have a significant association with readmission in the adjusted model (Figure 5b). However, there was weak evidence that regular transfers are associated with a minor increase in odds of readmission (OR = 1.02, 95%CI 1.00-1.04). Site transfers were associated with lower mortality (OR 0.72, 95%CI 0.64-0.80), with a decrease of 28% in odds of death for each unit increase in site transitions (Figure 5c). No relationship between atypical transfers or regular transfers and mortality was observed. Full multivariable regression models are given in supplementary Tables 3-5.

### Sensitivity analyses

The effect of atypical transfers was robust across the sensitivity analyses conducted, with results remaining consistent. Redefining atypical transfers by using the TFC to determine patient specialities reduced the number of patients undertaking at least one atypical transfer by 22.8% (n=5,473). Nevertheless, effect sizes remained consistent, with the magnitude of the impact of atypical transfers on LOS only marginally decreasing (AME = 2.58; 95%CI 2.28 – 2.87, p<0.001). Meanwhile, using an alternative categorisation of surgical procedures (See Supplementary Note 1c) increased the effect size to 3.03 days (95%CI 2.75-3.32, p<0.001) but attenuated the association between regular transfers and 30-day readmission (OR = 1.01; 95%CI 0.99-1.03, p=0.22). Removing influential outliers by large values of standardised residuals resulted in 149 observations being dropped from the LOS model, 19 from the mortality model and none from the readmission model but did not meaningfully change the main effects. All sensitivity analyses are presented in Supplementary Tables 6-8.

### Reasons underlying atypical transfers

Open coding identified 12 distinct reasons for atypical movements, classified under four main themes: ‘complex clinical journeys’, ‘non-clinical factors’, ‘a need for services and facilities’, and ‘unusual pathways’. The four themes are outlined if the following section. Table 3 details the codes and their descriptions, alongside illustrative quotations from the data.

**Table 3.**

| Code and description | Illustrative quotes |
| --- | --- |
| **Theme 1: Complex clinical journey** |  |
| **Co-occurring clinical needs**<br>The patient is moved between two different wards to receive input from a second speciality due to multimorbidity | “A lot of the renal patients that would have been there would also have been cardiology patients. Cause if you have a heart problem, you’re likely to have kidney problems and vice versa” – Interview 2, participant S12<br>“So you need to remember the...stroke unit ... the national guidelines state they need to stay a certain percentage of their hospital stay in the acute stroke unit... [there are] usually patients who move out from there because they don’t, no longer require stroke care or because they need another speciality for example vascular, or if that’s causing a stroke, they need an intervention or a cardiology issue is causing them a stroke, so they tend to not have [an] atypical transfer.” - Focus group 2, participant S07 |
| **Unexpected adverse event**<br>An unanticipated event results in a move to another speciality ward for treatment | “So the stroke units feeding into it could be anyone at anytime from A&E, or any of the inpatient wards can have a stroke while in hospital. So that could account for the transfers into them” – Interview 2, participant S12 |
| **Uncertainty over clinical speciality**<br>A transfer taking place due to ambiguity over the most appropriate clinical speciality for a patient | “Previously [the CDU] was used to move people in there who may need to have fluids, who may need to have something else...So it could be that they were just moved because they weren’t ready, or it was not clear what speciality they needed to go to... You know, and then sometimes we would have people that were in there for days, but it was just to see ... if we could get them home from there, rather than going into an acute bed.” – Focus group 3, participant B03 |
| **Theme 2: Non-clinical factors** |  |
| **Creating capacity**<br>The patient is moved to an unrelated speciality to create bed capacity for an incoming patient on the ward | “...[it] would be possible but unusual cause one is medicine or one is surgical so, yeah. I mean, it’s definitely possible, because the problem is we outline patients as well if there’s no capacity. So I’m guessing, you know, we’ve outlined orthopaedics and surgery on medicine.” – Interview 1, Participant S13<br>“Because out of hours, oncology even though the patient is known under oncology, if we don’t have any capacity at all on the oncology wards, we normally put patients or place patients on the medical wards. Depending on the condition of the patients, because some of the oncology patients [need] cardiac monitoring for the acute phase.” – Focus group 3, participant S11 |
| **Intra-divisional transfers**<br>A move to a speciality ward within the same overarching medical or surgical division to create bed capacity for an incoming patient | “If for example, if they ran out of beds and because we cover quite a big area, we can't close services, so we need to be operational for 24 hours... we need to create capacity in this area. So very often we will speak with the stroke reg [registrar] who can identify people who are not in their acute phase, and they will be going to Ward S. Saying this, you wouldn't, kind of, *you* would say outlier, but its not – as [participant] said before because they're staying in the same division which is neuroscience” – Focus group 3, participant S09 |
| **Admitting capability**<br>A temporarily placement on a ward out-of-hours | “Out of hours...for example, oncology, neurology, they don't admit patients. The patient is admitted under acute medicine and the speciality will take them over [the] next day” - Focus group 3, participant S09 |
| **Correcting an incorrect placement**<br>The patient is returned to their home ward following a transfer for creating capacity | “Some of the destinations of the atypical transfers aren't necessarily atypical destinations, they are the right destination, but they've gone from the wrong start point. So actually, its the ward they're going from is the atypical environment, and then actually we're almost correcting an incorrect placement. So if we put an orthopaedic patient on Ward L... that could actually be putting a patient back into the right place” - Focus group 1, participant S01 |
| Theme 3: Specific needs for services and facilities |  |
| **Rehabilitation services**<br>A transfer for the purposes of delivering rehabilitation services | “...a lot of our patients are admitted to the [acute medical unit] could be admitted with the fall from home, and where they need orthogeriatric input, even though they have fractures that don't need like surgical input, they might still need orthopaedic rehabilitation. So, we do get increasingly more patients on the [acute medical unit] that need the input that Ward J can offer them. So even though that's been flagged as a non-typical transfer, these days, I think more and more that's becoming a typical transfer because of [the] patients we're getting...[they] might be very elderly, not suitable for surgery. They might have like a broken arm and no one to look after them at home. So they need some rehab before they go home.” – Interview 2, participant S12<br>“...that's your stroke journey, you know, if you don't recover well and you need rehab you go to neuro rehab. So that kind of, that's kind of the stroke wards feed neuro rehab, you wouldn't get anybody going - people shouldn't really be going there from many other locations other than stroke” – Focus group 1, participant S01 |
| **Monitoring facilities**<br>The patient is moved to a ward which has cardiac monitoring equipment | “We don't have monitoring cardiac monitoring facilities in most...wards there apart from Ward J and ITU, so therefore if they become unwell, if they're needing a cardiac monitor then...there is the discussion between the medical team and then cardiology team, and maybe ... that would be an atypical transfer for them, because its not necessarily that the patient needs to be in the |
|  | cardiology wing...the patient could just been moved to a medical ward as well, with a cardiac monitoring facility.” – Focus group 2, participant S06<br>“... patients who become unwell and need an acute bed...and monitoring. I suppose it’s atypical, but necessary. You know, you need to do things sometimes for the safety of patients.” – Interview 3, participant S13 |
| **Side room availability**<br>The patient is moved to a ward with side room capacity for end-of-life care or to contain the spread of an infection | “... you mentioned about Ward Y. Cause obviously these are oncology patients, they tend to be neutropenic or end-of-life care, so they have quite limited side room available whereas Ward N has got 10 or 12 side rooms. So that’s why they tend to come to Ward N sometimes, they need a cardiac monitored side room, so that was clinically appropriate” – Focus group 2, participant S07<br>“...where you’ve got all specialities going in and not out, because the whole of ward is actually, they’re all side rooms, so where you have cases where people have developed diarrhoea, needing isolation immediately [its] usually the first port of call if they’re not requiring a monitored side room, so that’s where you probably see lots of the cases there” – Focus group 2, B01 |
| Theme 4: Unusual pathways |  |
| **Rare paths**<br>An obscure movement pattern with no clear explanation | “Participant S01: ...that’s a very very odd move. And I’ll be really interested to know what the others think might be a reason for that, cause I’ve been trying to think why we would move someone into CDU from a ward.<br>Participant B01: So I definitely think I’ve never done it [laugh] but do you think it, again could be just that there’s no beds and there just trying to move them, and they’re waiting for transport or something -<br>Participant S01: yeah, yeah, or we need a monitor somewhere<br>Participant B01: Yeah, someone waiting to go home so I’ll just put them into CDU for now, while they wait, yeah.” – Focus group 1, Participants S01 and B01 |
| **Exceptional events**<br>A transfer relating to an exceptional incident | “Participant S04: Another transfer or move of patient...operational wise is the trauma patients that involves police escort and so it’s basically not clinical but we move them to separate all the trauma patients, due too, I don’t know, gang members. So we can’t put them in one ward, so specifically they are supposed to be all in Trauma because they are all trauma patients, but due to the safety of patients and staff, they have to be separated in order to, operationally make wards and staff safe.<br>Participant S01: That’s a very good point. So we deliberately sometimes have to outlie people for the safety of the wards because we often have both sides of a fracas in the streets of London, both come into our A&E department; you can’t put them on the same ward because there will be a fight... one of them ends up being an atypical transfer”– Focus group 1, participant S04 and S01 |

#### Complex clinical journeys

Participants highlighted the possibility that some patients have complex clinical journeys, needing input from multiple specialities as their condition evolves. Co-occurring clinical needs could therefore result in transfers across wards with differing but inter-related specialities such as cardiology and renal wards, or stroke and vascular wards. Patients who suffered from unexpected adverse events such as a stroke or a fall may likewise require treatment across differing speciality wards equipped to treat their conditions. Participants also discussed a population of patients whose clinical complexity meant that the most appropriate speciality was unclear, resulting in multiple transfers as patients were assessed and their treatment course decided. This group included patients being transferred from clinical decisions units to in-hospital wards.

#### Non-clinical factors

A consistent explanation for atypical transfers offered across the data was the possibility that atypical transfers occur for operational reasons. Patients could be outlied across different divisions, such as medicine or surgery to create capacity for incoming patients. It is important to note that these transfers could not always be clearly distinguished from those of patients with complex journeys without additional patient-level information:

> Well two things, it could be typical or atypical. Typical if for example a patient is coming with nausea and vomiting, [who] has developed a surgical problem - then that would be a typical transfer, because … it’s not a medical problem anymore it’s a surgical…However, it becomes atypical if for example we’re inundated with quite a lot of medical patients in A&E… and they [surgical unit] have got something like 10 beds … that they don’t need… I would identify patients, with the help of the medical team, to outlie patients into the spaces. It’s because we need to deal with capacity issues and safety of A&E. - Focus group 2, Participant S06

Atypical transfer patients may also be moved within their overarching clinical division to create bed spaces. While such transfers could still be classed as non-clinical, participants typically distinguished these patients, who would remain in an appropriate overarching bed base, from strict outlier patients. One participant highlighted that some speciality wards are not able to admit patients out-of-hours, resulting in an intermediate general ward being used to admit the patient.

#### A need for services and facilities

Another theme across the data involved atypical transfers to a specific service or facility. These movements were distinct from complex clinical journeys in that they occurred in order for the patient to reach a specific service, equipment or infrastructure, as opposed to more broadly receive input from a second clinical speciality. A common example was the need for rehabilitation services, which were offered on specific wards rather than as a mobile service. In addition, cardiac monitoring was a scarce resource and usually required the patient to be moved to a ward which provided this equipment. Similarly, if a patient became infected on an open bay ward, a transfer would be required to an isolated single side-room, which are only available in a minority of wards.

#### Unusual pathways

Finally, the ‘unusual pathways’ theme comprises a small number of atypical transfers which participants highlighted as obscure, and possibly due to exceptional reasons. Unusual reasons included major incidents, ward closures, and circumstances such as two individuals being involved in a confrontation being admitted to the same trauma ward.

## DISCUSSION

### Principal findings

An increasing evidence base associates intrahospital transfers with adverse outcomes. However, our analysis shows that not all ward transfers are equal. In a large-scale retrospective cohort study, we show the feasibility of a novel data-driven approach, which leverages rich EHR data to characterise atypical transfers. We found that among hospitalised patients who undertake at least one transfer during their spell, 12.9% use ‘atypical’ routes between uncommon pairs of wards for their speciality. Such patients experience an approximate 2.8 day increase in LOS, after controlling for regular transfers, site transfers and case-mix variables. The effect size was 0.9 days larger than that of a regular intrahospital transfer to or from any other ward in the hospital (AME = 1.92; 95%CI 1.82-2.03). No relationships between atypical transfers and mortality or 30-day readmission were observed. A secondary finding is that different types of patient movement have differing effects. Transfers between hospital sites within one trust were associated with a 28% reduction in odds of mortality (OR = 0.72; 95%CI 0.64-0.80), and regular transfers with a small increase in odds of readmission (OR = 1.02; 95%CI 1.00-1.04). Box 1 is an illustrative fictitious example of an atypical patient’s hospital journey, based on true trajectories.

#### Box 1: Example atypical patient

An elderly patient was admitted to the acute medical unit at 3am over a weekend, with influenza. After 2 days of being treated by a general medicine consultant, they were moved during the night to a surgical ward, which typically treats trauma and neurosurgery patients. They remained on the ward another day before being transferred to a medical ward which usually sees general medicine, genitourinary medicine and respiratory medicine patients before being discharged. These two movements were deemed atypical because the wards have no overlapping specialities. The patient was in hospital for 10 days.

Qualitative focus groups and interviews with experienced site nurse practitioners and bed managers revealed four overarching themes surrounding the decision making underlying atypical transfers: complex patient journeys, meaning the patient did not fit into any common groups of services (theme 1), a lack of capacity resulting in non-clinical transfers within and across overarching major divisions (theme 2), or a particular clinical need triggering a transfer to reach specific services and facilities (theme 3). More exceptionally, some transfers had no clear explanation, and possibly arose because of a rare, unplanned event (theme 4).

### Triangulation and interpretation in light of other evidence

While our study did not directly explore the effect of placing patients on clinically inappropriate wards (and excluded patients staying only on one ward), its findings are consistent with several studies on the impact of outlying patients, which report increases in LOS by 0.1 to 1 days.[18], [19], [42] Only one quantitative study has explored the topic of outlying patients using UK hospital data, reporting an increase in LOS but no effect on mortality.[18] The qualitative strand of the study suggests our approach captured ‘partial outliers’ who move between appropriate and inappropriate wards, or vice versa (theme 2). Quantitatively, we found that patients experiencing atypical transfers were more likely to be admitted out-of-hours, which has been associated with outlying status in other studies.[43] The effect of undergoing an atypical transfer cannot be separated from the subsequent impact of being treated on a potentially inappropriate ward, which may account for increased LOS. However, as clinical factors were also suggested as reasons underlying the transfers, it is not the sole explanation of the effect. Therefore, while atypical transfers have a comparable effect on LOS to outlying patients, they do not necessarily imply that the transfer is clinically inappropriate.[18]

The qualitative component of the study highlighted two clinical reasons which may lead to atypical transfers. The patient does not equivocally fit into commonly paired wards (theme 1), or the patient needed access to a specific service or facility (theme 3) such as the need for cardiac monitoring paired with an orthopaedic condition, where the nursing skills to look after both factors is seldom co-located in one ward. The quantitative strand of this study also supports these explanations. Patients taking atypical routes were more likely to have multiple primary ICD-10 codes during their spell, potentially reflecting a complex clinical condition and generally experienced more procedures. The need for isolation due to infection was discussed as a reason behind atypical transfers qualitatively, but quantitative findings showed that removing infected patients from the analysis did not attenuate the association with increased LOS, making infection control an unlikely driver of increased LOS. In other literature, cases of population-capacity misalignment comparable to the first theme have been described in qualitative work.[44]–[46] Kreindler *et al.,* highlight the complexities faced by hospital managers in Canada when patients have significant co-occurring needs, such as dementia and pneumonia.[47] The patient is then moved, introducing them to a new team and extending their spell. Atypical routes also frequently involved observation units, which have been associated with ad-hoc use where a lack of alternative pathways exist.[41], [48]

An important distinction our study has made to previous literature is that it is exploring the whole patient movement history, rather than a single location. This approach highlights the outcomes of patients who experience transfers. Ward transfers are complex procedures, both in the physical process required to move a patient, which can be destabilising,[49] and the decision-making processes behind them. Transfers are a vulnerable time for patients and can leave them feeling anxious, disorientated, and ‘forgotten’ by staff,[50], [51] particularly on an inappropriate ward.[44] Transfers also require cooperation, negotiation, and trust between the sending and receiving clinicians, which is strengthened by familiarity.[52], [53],[54] Clinical handovers can be prone to workarounds and communication breakdowns even within one clinical team,[55] and exacerbated when occurring across units, specialities and physical boundaries.[56] Patient movement may therefore be a potential additional driver behind increased LOS, which should be considered in studies exploring patient hospital locations.

It is also important to note that among patients with at least one transfer, each additional regular transfer increased LOS by approximately 1.9 days, suggesting that even those undertaking regular transfers experience an associated increase in LOS, after adjusting for case-mix factors. However, this relationship may differ when considering patients unexposed to movement. Others have reported large increases in LOS after intrahospital transfers,[11], [57] highlighting the importance of avoiding transfers where possible. Similarly, while the outcomes of patients following an inter-hospital transfer have been studied,[58], [59] the impact of a transfer to a hospital within a single trust has been unexplored. Our finding that such transfers are associated with a decrease in mortality suggests that localisation of specialist and general hospitals within a trust is a successful model of care, in the context of a large urban hospital trust. Our models controlled for the patient acuity variables available, however it is possible that only patients most likely to survive are transferred between sites, leading to residual confounding. The differences between NHS trusts which co-locate their services in a single site, versus geographically dispersed sites are areas of possible future investigation.

### Strengths and limitations

A key strength of our study is the use of a data directed definition of ward specialities and atypical transfers, coupled with a qualitative exploration of their meaning. The data driven definition captures the functional use of wards, rather than a pre-defined, theoretical use. This is an important strength, as the boundaries of a specific service can become blurred in the day-to-day running of a hospital, particularly as bed pools shift over time, leading to possible misclassification of outlying patients.[60] The task of matching specialities to patient needs is highly complex and organisation dependent, and pre-defining ward specialities may also overlook the fact that staff treating many outliers can become as familiar in caring for them as an inlier patient. With some studies reporting as many as 40% of the patient population to be outliers,[43] the causal hypothesis that outlier patients have increased adverse outcomes because they are treated by a nursing team which is inexperienced with their condition may not hold. Nevertheless, using a heuristic also draws arbitrary cut off points, and speciality definitions vary depending on the chosen TFC, which can also be subject to recording inconsistencies. While this limitation is mitigated by the large study size, the fact that we removed erroneous spells on several criteria, and that consistent results were obtained when using the first and last patient TFC, inaccuracies and inconsistencies are a known limitation to retrospective EHR data analyses, which should be considered when interpreting results.[61] Importantly, the qualitative strand of this study addresses some of the weaknesses of the quantitative strand, by verifying ward specialities and explaining the possible purposes underlying atypical transfers. Moreover, our quantitative method can be applied to other routinely collected datasets for validation without the considerable domain knowledge needed to allocate ward specialities manually. It is also possible that atypical transfers are a marker of health system strain akin to outlying patients, leading to residual confounding from broader health system factors such as understaffing, which was not directly adjusted. While it is difficult to attribute causality between this exposure and the environment in which it occurs, adjusting for weekend/out-of-hours admissions when hospitals typically function with reduced staffing, as well as approximate seasonality did not account for the effect on LOS.

It is also important to consider that the specific reasoning behind transfers cannot be systematically analysed without patient notes, and that the qualitative data may be limited by a small sample size. However, the qualitative strand had a narrow aim, and the participants recruited held a large amount of knowledge and depth of experience relevant to this aim. The principles of information power suggests such a study does not require a large sample size.[62]

Finally, the generalisability of our findings is unclear. Three other studies have used network analysis to explore patient transfers within hospitals and demonstrated that rare, low frequency transfers also occur in these hospital environments,[30], [63], [64]. This suggests atypical transfers (with respect to frequency) are not isolated to our setting; however, authors did not link these to patient outcomes.

### Implications

Our findings have implications for hospital design and future research. Firstly, we have demonstrated the feasibility of a data-driven method to identify patients which, for any reason, move between uncommon pairs of services using EHR data. These initial findings support further exploration of ward movements, as well as the potential for hospital trusts to leverage their own EHRs for optimising patient pathways in real-time. For example, the identification of complex patients though atypical movements could guide the creation of multi-condition services based on clusters of co-occurring needs.[47] Models of population segmentation are an important complement to the shift from single conditions to integrated, needs-based care systems,[65], [66] and can be supported by data-driven methods.[67]–[69] While focus has been given to primary care interventions,[65] population segmentation interventions in secondary care may improve hospital flow through the introduction of integrated units which have fluid resources and wide eligibility criteria, to better accommodate patients with a ‘shifting fuzzy set of needs’.[47] Atypical movements can be a potential system focused metric used alongside others to develop segmentation logic. As the UK government looks to expand hospital infrastructure,[70] such analyses have a place in informing policy on the medical built environment, in combination with knowledge from clinicians, hospital managers and healthcare architects.

Secondly, when patients must be moved due to non-clinical reasons, our analysis also suggests that transfers to wards with a similar speciality profile reduces subsequent LOS. In the highly complex, non-linear hospital system, it is important to consider the downstream effects of policies which aim to rapidly decant the ED.[71], [72] These can result in more patients placed on wards with any available space, increasing atypical transfers.[73] While our findings show that atypical transfers do not increase mortality or readmission, their relationship to increased LOS suggests that such strategies may exacerbate exit-block in the long-term, as patients remain in hospital for longer. From a systems perspective, minimising atypical transfers helps to sever the cycle of bed-blocking that occurs when patients on the wrong ward spend extra days in hospital, thereby further diminishing access to beds.[74] However, future work is needed to understand the generalisability of these findings, given the heterogeneity of ward management practices across hospitals. It is also important to consider that a suboptimal patient transfer for one patient may be crucial for the care of the individual taking their bed space. Qualitative exploration to elucidate the challenges that bed managers face, around which literature is limited.[75] Our analysis therefore provides a starting point for identifying clusters of patients who have moved between unexpected pairs of wards, with a view to optimise pathways for future patients.

## Conclusion

Routinely collected EHR data give us the opportunity to examine the true hospital usage and evaluate deviant patient journeys, which may otherwise go undetected. Atypical ward transfers are associated with a significant increase in the patient’s LOS, which is detrimental to both the individual and the wider health system. The physical movement, unfamiliarity between services, and treatment of the patient on a potentially mismatched ward may be factors contributing to this effect. The causes of atypical transfers, and the broader impact of patient movement must be better understood and considered in hospital policy and design. Our work provides an important first step in identifying unusual patient movement and its impacts.

## Data Availability

Data may be obtained from a third party and are not publicly available. Deidentified patient data cannot be made publicly available due to information governance restrictions. Access to the data sets used in this paper via a secure environment will be reviewed on request by Imperial College Healthcare NHS Trust.

## Acknowledgements

This research has been completed using the National Institute for Health Research Health Informatics Collaborative (NIHR HIC) data resources and supported by the Imperial Clinical Analytics Research and Evaluation (iCARE) team. Data management (and analytical support) was provided by the Big Data and Analytical Unit (BDAU) at the Institute of Global Health Innovation (IGHI). Imperial College London is grateful for the support from the North West London NIHR Applied Research Collaboration.

We thank Dr Erik Mayer and Dr Benjamin Post for the helpful discussions on the data analysis and clinical interpretation of the findings.

## Author Contributions

EM, PE and CC conceptualised the research question. EM, overseen by PE and CC, conducted the data analysis. EM wrote and revised the manuscript. LM conducted the data curation and anonymisation KH, AB, RK, provided expert advice and critical review of the paper prior to submission. The corresponding author attests that all listed authors meet authorship criteria and that no others meeting the criteria have been omitted.

## Disclaimer

This article presents independent research commissioned by the National Institute for Health Research (NIHR) under the Applied Research Collaboration (ARC) programme for Northwest London. The views expressed in this publication are those of the author(s) and not necessarily those of the NHS, the NIHR or the Department of Health and Social Care.

