## Supplementary Information for "The impact of atypical intrahospital transfers on patient outcomes: a mixed methods study"

### Univariable regression results:

**Table 1a:** Unadjusted average marginal effects for the relationship between atypical transfers, regular transfers, site transfers and increased length of stay derived from a gamma GLM with log link.

| Variable | AME | Lower 95% CI | Upper 95% CI | p-value |
|---|---|---|---|---|
| Atypical transfers | 3.34 | 3.06 | 3.68 | <0.001 |
| Regular transfers | 3.09 | 2.99 | 3.20 | <0.001 |
| Site transfers | 7.70 | 6.11 | 9.30 | <0.001 |

**Table 1b:** Unadjusted odds ratios (OR) for the relationship between atypical, routine and site transfers and in-hospital mortality derived from logistic regressions.

| Variable | OR | Lower 95% CI | Upper 95% CI | p-value |
|---|---|---|---|---|
| Atypical transfers | 1.14 | 1.05 | 1.23 | 0.001 |
| Regular transfers | 1.18 | 1.16 | 1.20 | 0.001 |
| Site transfers | 1.00 | 0.91 | 1.09 | 0.98 |

**Table 1c:** Unadjusted odds ratios (OR) for the relationship between atypical, routine and site transfers and 30-day emergency readmission derived from logistic regressions.

| Variable | OR | Lower 95% CI | Upper 95% CI | p-value |
|---|---|---|---|---|
| Atypical transfers | 0.98 | 0.93 | 1.03 | 0.37 |
| Regular transfers | 1.02 | 1.00 | 1.03 | 0.02 |
| Site transfers | 1.02 | 0.96 | 1.08 | 0.51 |

**Table 2: Full table of patient characteristics**

| | No atypical transfers (N=48064) | At least ones atypical transfer(N=7088) | Total (N=55152) |
|---|---|---|---|
| Routine ward transfers | | | |
| **Median** | 1 | 0 | 1 |
| **Q1, Q3** | 1, 2 | 0, 2 | 1, 2. |
| Site transfers | | | |
| **Median** | 0 | 0 | 0 |
| **Q1, Q3** | 0,0 | 0, 0 | 0, 0 |
| Age (years) | | | |
| **18-39** | 6273 (13.1%) | 906 (12.8%) | 7179 (13.0%) |
| **40-65** | 15332 (31.9%) | 2205 (31.1%) | 17537 (31.8%) |
| **over 65** | 26459 (55.0%) | 3977 (56.1%) | 30436 (55.2%) |
| Gender | | | |
| **Female** | 22419 (46.6%) | 3589 (50.6%) | 26008 (47.2%) |
| **Male** | 25645 (53.4%) | 3499 (49.4%) | 29144 (52.8%) |
| Length of stay (days) | | | |
| **Median** | 7 | 9 | 7.2 |
| **Q1, Q3** | 3.3, 14.4 | 4.2, 19.6 | 3.5, 14.9 |
| Elixhauser comorbidities (n, %) | | | |
| **>=5** | 9219 (19.2%) | 1338 (18.9%) | 10557 (19.1%) |
| **0** | 5254 (10.9%) | 839 (11.8%) | 6093 (11.0%) |
| **1-4** | 33591(69.9%) | 4911 (69.3%) | 38502 (69.8%) |
| Discharge destination | | | |
| **Not usual place of residence** | 7190 (15.0%) | 1212 (17.1%) | 8402 (15.2%) |
| **Usual place of residence** | 40874 (85.0%) | 5876 (82.9%) | 46750 (84.8%) |
| Primary diagnosis count (n, %) | | | |
| **Mean (SD)** | 1.2 (0.4) | 1.3 (0.5) | 1.2 (0.4) |
| **Range** | 1.0 - 9.0 | 1.0 - 8.0 | 1.0 - 9.0 |
| Intensive Care Admission | | | |
| **0** | 44747 (93.1%) | 6678 (94.2%) | 51425 (93.2%) |
| **1** | 3317 (6.9%) | 410 (5.8%) | 3727 (6.8%) |
| mortality | | | |
| **0** | 45454 (94.6%) | 6676 (94.2%) | 52130 (94.5%) |
| **1** | 2610 (5.4%) | 412 (5.8%) | 3022 (5.5%) |
| Weekend admission | | | |
| **FALSE** | 36749 (76.5%) | 5294 (74.7%) | 42043 (76.2%) |
| **TRUE** | 11315 (23.5%) | 1794 (25.3%) | 13109 (23.8%) |
| Out of hours admission (7pm-7am) | | | |
| **0** | 25273 (52.6%) | 3400 (48.0%) | 28673 (52.0%) |
| **1** | 22791 (47.4%) | 3688 (52.0%) | 26479 (48.0%) |
| Diagnostic imaging procedures (n, %) | | | |
| **0** | 23701 (49.3%) | 2646 (37.3%) | 26347 (47.8%) |
| **0-3** | 22206 (46.2%) | 3891 (54.9%) | 26097 (47.3%) |
| **4-6** | 1835 (3.8%) | 440 (6.2%) | 2275 (4.1%) |
| **Over 6** | 322 (0.7%) | 111 (1.6%) | 433 (0.8%) |
| Minor surgical procedures (n, %) | | | |
| **0** | 40339 (83.9%) | 5881 (83.0%) | 46220 (83.8%) |
| **1-2** | 7193 (15.0%) | 1103 (15.6%) | 8296 (15.0%) |

| | | | |
|---|---|---|---|
| **3-5** | 473 (1.0%) | 87 (1.2%) | 560 (1.0%) |
| **Over 5** | 59 (0.1%) | 17 (0.2%) | 76 (0.1%) |
| Major surgical procedures (n, %) | | | |
| **0** | 41334 (86.0%) | 5857 (82.6%) | 47191.0 (85.6%) |
| **1-3** | 6198 (12.9%) | 1168 (16.5%) | 7366.0 (13.4%) |
| **4-6** | 453 (0.9%) | 55 (0.8%) | 508.0 (0.9%) |
| **Over 6** | 79 (0.2%) | 8 (0.1%) | 87.0 (0.2%) |
| Ethnicity code description (n, %) | | | |
| **African** | 1816 (3.8%) | 243 (3.4%) | 2059.0 (3.7%) |
| **Any other Asian background** | 2021 (4.2%) | 277 (3.9%) | 2298.0 (4.2%) |
| **Any other Black background** | 998 (2.1%) | 159 (2.2%) | 1157.0 (2.1%) |
| **Any other ethnic group** | 6691 (13.9%) | 1013 (14.3%) | 7704.0 (14.0%) |
| **Any other mixed background** | 289 (0.6%) | 53 (0.7%) | 342.0 (0.6%) |
| **Any other White background** | 5629 (11.7%) | 843 (11.9%) | 6472.0 (11.7%) |
| **Bangladeshi** | 267 (0.6%) | 37 (0.5%) | 304.0 (0.6%) |
| **British** | 16371 (34.1%) | 2414 (34.1%) | 18785.0 (34.1%) |
| **Caribbean** | 2366 (4.9%) | 368 (5.2%) | 2734.0 (5.0%) |
| **Chinese** | 235 (0.5%) | 28 (0.4%) | 263.0 (0.5%) |
| **Indian** | 2093 (4.4%) | 262 (3.7%) | 2355.0 (4.3%) |
| **Irish** | 1841 (3.8%) | 285 (4.0%) | 2126.0 (3.9%) |
| **Pakistani** | 474 (1.0%) | 66 (0.9%) | 540.0 (1.0%) |
| **Unknown** | 6555 (13.6%) | 980 (13.8%) | 7535.0 (13.7%) |
| **White and Asian** | 114 (0.2%) | 15 (0.2%) | 129.0 (0.2%) |
| **White and Black African** | 82 (0.2%) | 16 (0.2%) | 98.0 (0.2%) |
| **White and Black Caribbean** | 222 (0.5%) | 29 (0.4%) | 251.0 (0.5%) |
| Major diagnostic category (n, %) | | | |
| **Certain infectious and parasitic diseases** | 3371 (7.0%) | 420 (5.9%) | 3791 (6.9%) |
| **Congenital malformations, deformations and chromosomal abnormalities** | 26 (0.1%) | 5 (0.1%) | 31 (0.1%) |
| **Diseases of the blood and blood-forming organs and certain disorders involving the immune mechanism** | 1023 (2.1%) | 91 (1.3%) | 1114 (2.0%) |
| **Diseases of the circulatory system** | 8985 (18.7%) | 1253 (17.7%) | 10238 (18.6%) |
| **Diseases of the digestive system** | 4223 (8.8%) | 543 (7.7%) | 4766 (8.6%) |
| **Diseases of the ear and mastoid process** | 54 (0.1%) | 19 (0.3%) | 73 (0.1%) |
| **Diseases of the eye and adnexa** | 147 (0.3%) | 33 (0.5%) | 180 (0.3%) |
| **Diseases of the genitourinary system** | 4095 (8.5%) | 575 (8.1%) | 4670 (8.5%) |
| **Diseases of the musculoskeletal system and connective tissue** | 1006 (2.1%) | 281 (4.0%) | 1287 (2.3%) |
| **Diseases of the nervous system** | 1114 (2.3%) | 297 (4.2%) | 1411 (2.6%) |
| **Diseases of the respiratory system** | 7717 (16.1%) | 859 (12.1%) | 8576 (15.5%) |
| **Diseases of the skin and subcutaneous tissue** | 906 (1.9%) | 136 (1.9%) | 1042 (1.9%) |
| **Endocrine, nutritional and metabolic diseases** | 1262 (2.6%) | 166 (2.3%) | 1428 (2.6%) |

| | | | |
|---|---|---|---|
| **External causes of morbidity** | 5 (0.0%) | 0 (0.0%) | 5 (0.0%) |
| **Factors influencing health status and contact with health services** | 129 (0.3%) | 18 (0.3%) | 147 (0.3%) |
| **Injury, poisoning and certain other consequences of external causes** | 6018 (12.5%) | 751 (10.6%) | 6769 (12.3%) |
| **Mental, Behavioral and Neurodevelopmental disorders** | 111 (0.2%) | 22 (0.3%) | 133 (0.2%) |
| **Neoplasms** | 2722 (5.7%) | 654 (9.2%) | 3376 (6.1%) |
| **Symptoms, signs and abnormal clinical and laboratory findings, not elsewhere classified** | 5150 (10.7%) | 965 (13.6%) | 6115 (11.1%) |

**Full multivariable regression results**

**Table 3:** Multivariable generalised linear model (GLM) regression with a gamma distribution and log link predicting hospital length of stay (n = 55152). Results are presented as adjusted average marginal effects in days, with 95% CI based on standard errors clustered at the patient level (n = 38101).

| variable | estimate | SE | lower 95% CI | upper 95% CI | p-value |
|---|---|---|---|---|---|
| (Intercept) | 8.26 | 0.03 | 7.74 | 8.8 | <0.001 |
| Atypical transfers | 1.22 | 0.01 | 1.19 | 1.24 | <0.001 |
| Regular transfers | 1.14 | 0 | 1.13 | 1.15 | <0.001 |
| Site transfers | 1.23 | 0.01 | 1.21 | 1.26 | <0.001 |
| *Reference aged 18 to 40 years* | | | | | |
| 40-65 | 1.17 | 0.01 | 1.14 | 1.2 | <0.001 |
| Over 65 | 1.53 | 0.01 | 1.49 | 1.57 | <0.001 |
| *Reference Over 5 Elixhauser comorbidities* | | | | | |
| factor(index)0 | 0.68 | 0.02 | 0.66 | 0.71 | <0.001 |
| 1-4 | 0.85 | 0.01 | 0.84 | 0.87 | <0.001 |
| *Reference not weekend admission* | | | | | |
| Weekend admission | 1.01 | 0.01 | 0.99 | 1.03 | 0.16 |
| *Reference Discharge not to usual place of residence* | | | | | |
| Usual place of residence | 0.64 | 0.01 | 0.63 | 0.66 | <0.001 |
| *Reference no ICU attendance* | | | | | |
| ICU admission | 1.17 | 0.02 | 1.13 | 1.22 | <0.001 |
| *Reference female* | | | | | |
| Male | 0.97 | 0.01 | 0.95 | 0.99 | <0.001 |
| *Reference 1 primary diagnosis* | | | | | |
| 2-3 | 1.43 | 0.01 | 1.4 | 1.47 | <0.001 |
| Over 3 | 2.29 | 0.08 | 1.96 | 2.66 | <0.001 |
| *Reference 0 intermediate surgical procedures* | | | | | |
| 1-3 | 1.62 | 0.01 | 1.6 | 1.65 | <0.001 |
| 4-6 | 2.85 | 0.02 | 2.73 | 2.98 | <0.001 |
| Over 6 | 3.69 | 0.05 | 3.35 | 4.05 | <0.001 |
| *Reference 0 minor surgical procedures* | | | | | |
| 1-2 | 1.12 | 0.01 | 1.09 | 1.15 | <0.001 |
| 3-5 | 1.35 | 0.04 | 1.24 | 1.46 | <0.001 |
| Over 5 | 1.37 | 0.11 | 1.1 | 1.7 | <0.001 |
| *Reference 0 intermediate surgical procedures* | | | | | |
| 1-4 | 1.07 | 0.01 | 1.05 | 1.09 | <0.001 |
| 5-7 | 1.53 | 0.05 | 1.4 | 1.67 | <0.001 |
| 8-10 | 1.58 | 0.06 | 1.4 | 1.79 | <0.001 |
| Over 10 | 2.53 | 0.09 | 2.1 | 3.05 | <0.001 |

| | | | | | |
|---|---|---|---|---|---|
| *Reference 0 major surgical procedures* | | | | | |
| 1-3 | 1.37 | 0.01 | 1.34 | 1.41 | <0.001 |
| 4-6 | 1.77 | 0.04 | 1.62 | 1.93 | <0.001 |
| Over 6 | 2.17 | 0.1 | 1.77 | 2.66 | <0.001 |
| *Reference admission between 7am and 7pm* | | | | | |
| Out-of-hours admission | 1.06 | 0.01 | 1.04 | 1.08 | <0.001 |
| *Reference African ethnicity code* | | | | | |
| Any other Asian background | 0.85 | 0.03 | 0.8 | 0.9 | <0.001 |
| Any other Black background | 0.94 | 0.03 | 0.88 | 1.01 | 0.09 |
| Any other ethnic group | 0.87 | 0.02 | 0.83 | 0.91 | <0.001 |
| Any other mixed background | 0.82 | 0.06 | 0.73 | 0.91 | <0.001 |
| Any other White background | 0.86 | 0.02 | 0.82 | 0.91 | <0.001 |
| Bangladeshi | 0.82 | 0.06 | 0.73 | 0.91 | <0.001 |
| British | 0.92 | 0.02 | 0.88 | 0.96 | <0.001 |
| Caribbean | 0.98 | 0.03 | 0.93 | 1.04 | 0.58 |
| Chinese | 0.88 | 0.06 | 0.78 | 0.99 | 0.04 |
| Indian | 0.91 | 0.03 | 0.86 | 0.97 | <0.001 |
| Irish | 0.94 | 0.03 | 0.88 | 0.99 | 0.03 |
| Pakistani | 0.85 | 0.05 | 0.77 | 0.93 | <0.001 |
| Unknown | 0.87 | 0.02 | 0.83 | 0.92 | <0.001 |
| White and Asian | 0.75 | 0.09 | 0.63 | 0.88 | <0.001 |
| White and Black African | 0.81 | 0.1 | 0.67 | 0.98 | 0.03 |
| White and Black Caribbean | 0.95 | 0.06 | 0.84 | 1.08 | 0.43 |
| *Reference certain infectious and parasitic diseases ICD-10 chapter* | | | | | |
| Congenital malformations, deformations and chromosomal abnormalities | 0.83 | 0.17 | 0.6 | 1.17 | 0.29 |
| Diseases of the blood and blood-forming organs and certain disorders involving the immune mechanism | 1.07 | 0.03 | 1 | 1.14 | 0.05 |
| Diseases of the circulatory system | 0.71 | 0.02 | 0.68 | 0.73 | <0.001 |
| Diseases of the digestive system | 0.68 | 0.02 | 0.65 | 0.71 | <0.001 |
| Diseases of the ear and mastoid process | 0.61 | 0.11 | 0.49 | 0.76 | <0.001 |
| Diseases of the eye and adnexa | 0.5 | 0.07 | 0.43 | 0.58 | <0.001 |
| Diseases of the genitourinary system | 0.97 | 0.02 | 0.93 | 1.01 | 0.11 |
| Diseases of the musculoskeletal system and connective tissue | 0.95 | 0.03 | 0.89 | 1.01 | 0.08 |
| Diseases of the nervous system | 0.91 | 0.03 | 0.85 | 0.96 | <0.001 |
| Diseases of the respiratory system | 0.92 | 0.02 | 0.89 | 0.96 | <0.001 |

| | | | | | |
|---|---|---|---|---|---|
| Diseases of the skin and subcutaneous tissue | 1.05 | 0.03 | 0.98 | 1.12 | 0.16 |
| Endocrine, nutritional and metabolic diseases | 0.81 | 0.03 | 0.76 | 0.86 | <0.001 |
| factor(chapter)External causes of morbidity | 0.94 | 0.42 | 0.41 | 2.17 | 0.89 |
| Factors influencing health status and contact with health services | 0.69 | 0.08 | 0.59 | 0.81 | <0.001 |
| Injury, poisoning and certain other consequences of external causes | 0.91 | 0.02 | 0.87 | 0.95 | <0.001 |
| Mental, Behavioral and Neurodevelopmental disorders | 1.02 | 0.08 | 0.86 | 1.2 | 0.85 |
| Neoplasms | 0.77 | 0.02 | 0.74 | 0.81 | <0.001 |
| Symptoms, signs and abnormal clinical and laboratory findings, not elsewhere classified | 0.68 | 0.02 | 0.65 | 0.71 | <0.001 |

**Table 4:** Multivariable logistic regression predicting in-hospital mortality (n = 55000). Results are presented as adjusted odds ratios, with 95% CI based on standard errors clustered at the patient level.

| **variable** | **estimate** | **SE** | **lower 95% CI** | **upper 95% CI** | **p-value** |
|---|---|---|---|---|---|
| (Intercept) | 0 | 0.24 | 0 | 0.01 | <0.001 |
| Atypical transfers | 0.97 | 0.04 | 0.89 | 1.05 | 0.41 |
| Regular transfers | 0.99 | 0.01 | 0.96 | 1.01 | 0.37 |
| Site transfers | 0.72 | 0.06 | 0.64 | 0.80 | <0.001 |
| *Reference aged 18 to 40 years* | | | | | |
| 40-65 | 3.26 | 0.16 | 2.38 | 4.46 | <0.001 |
| Over 65 | 8.61 | 0.16 | 6.33 | 11.71 | <0.001 |
| *Reference 0 Elixhauser comorbidities* | | | | | |
| Over 5 | 3.92 | 0.14 | 2.98 | 5.16 | <0.001 |
| 1-4 | 1.87 | 0.14 | 1.43 | 2.45 | <0.001 |
| *Reference not weekend admission* | | | | | |
| Weekend admission | 1.08 | 0.05 | 0.99 | 1.19 | 0.08 |
| *Reference female* | | | | | |
| Male | 0.98 | 0.04 | 0.9 | 1.06 | 0.55 |
| *Reference 1 primary diagnosis* | | | | | |
| ICU admission | 5.19 | 0.06 | 4.61 | 5.84 | <0.001 |
| *Reference 1 primary diagnosis* | | | | | |
| 2-3 | 2.6 | 0.05 | 2.36 | 2.87 | <0.001 |
| Over 3 | 3.23 | 0.25 | 1.99 | 5.26 | <0.001 |
| *Reference 0 imaging surgical procedures* | | | | | |

| | | | | | |
|---|---|---|---|---|---|
| 1-3 | 1.34 | 0.04 | 1.23 | 1.47 | <0.001 |
| 4-6 | 1.87 | 0.09 | 1.58 | 2.22 | <0.001 |
| Over 6 | 2.66 | 0.16 | 1.96 | 3.61 | <0.001 |
| *Reference 0 minor surgical procedures* | | | | | |
| 1-2 | 0.96 | 0.06 | 0.86 | 1.08 | 0.54 |
| 3-5 | 1.11 | 0.16 | 0.81 | 1.51 | 0.51 |
| Over 5 | 2.09 | 0.32 | 1.11 | 3.95 | 0.02 |
| *Reference 0 intermediate surgical procedures* | | | | | |
| 1-4 | 1.09 | 0.05 | 0.99 | 1.2 | 0.08 |
| 5-7 | 1.34 | 0.19 | 0.92 | 1.96 | 0.13 |
| 8-10 | 1.1 | 0.3 | 0.61 | 2 | 0.75 |
| Over 10 | 1.94 | 0.43 | 0.84 | 4.5 | 0.12 |
| *Reference 0 major surgical procedures* | | | | | |
| 1-3 | 0.71 | 0.07 | 0.62 | 0.81 | <0.001 |
| 4-6 | 0.5 | 0.19 | 0.34 | 0.73 | <0.001 |
| Over 6 | 0.42 | 0.44 | 0.18 | 1 | 0.05 |
| *Reference admission between 7am and 7pm* | | | | | |
| Out-of-hours admission | 1.1 | 0.04 | 1.01 | 1.19 | 0.02 |
| *Reference African ethnicity code* | | | | | |
| Any other Asian background | 1.21 | 0.16 | 0.89 | 1.64 | 0.23 |
| Any other Black background | 0.82 | 0.21 | 0.54 | 1.24 | 0.35 |
| Any other ethnic group | 1.08 | 0.14 | 0.83 | 1.41 | 0.55 |
| Any other mixed background | 0.84 | 0.33 | 0.44 | 1.58 | 0.59 |
| Any other White background | 1.18 | 0.14 | 0.9 | 1.54 | 0.23 |
| Bangladeshi | 0.66 | 0.34 | 0.34 | 1.29 | 0.23 |
| British | 1.14 | 0.13 | 0.89 | 1.47 | 0.29 |
| Caribbean | 1.04 | 0.15 | 0.77 | 1.4 | 0.81 |
| Chinese | 1.18 | 0.29 | 0.66 | 2.08 | 0.58 |
| Indian | 1.04 | 0.16 | 0.77 | 1.41 | 0.81 |
| Irish | 1.07 | 0.15 | 0.79 | 1.44 | 0.68 |
| Pakistani | 1.07 | 0.25 | 0.66 | 1.74 | 0.77 |
| Unknown | 1.15 | 0.13 | 0.88 | 1.49 | 0.3 |
| White and Asian | 0.81 | 0.46 | 0.33 | 1.99 | 0.65 |
| White and Black African | 1.16 | 0.61 | 0.35 | 3.85 | 0.81 |
| White and Black Caribbean | 0.69 | 0.42 | 0.3 | 1.57 | 0.38 |
| *Reference certain infectious and parasitic diseases ICD-10 chapter* | | | | | |
| Congenital malformations, deformations and chromosomal abnormalities | 1.82 | 1.04 | 0.24 | 13.95 | 0.56 |
| Diseases of the blood and blood-forming organs and certain disorders involving the immune mechanism | 0.31 | 0.26 | 0.19 | 0.53 | <0.001 |

| | | | | | |
|---|---|---|---|---|---|
| Diseases of the circulatory system | 0.63 | 0.07 | 0.54 | 0.73 | <0.001 |
| Diseases of the digestive system | 0.55 | 0.1 | 0.45 | 0.66 | <0.001 |
| Diseases of the ear and mastoid process | 0.19 | 1.02 | 0.03 | 1.38 | 0.1 |
| Diseases of the eye and adnexa | 0.16 | 0.77 | 0.03 | 0.7 | 0.02 |
| Diseases of the genitourinary system | 0.37 | 0.1 | 0.3 | 0.46 | <0.001 |
| Diseases of the musculoskeletal system and connective tissue | 0.28 | 0.19 | 0.19 | 0.4 | <0.001 |
| Diseases of the nervous system | 0.53 | 0.15 | 0.4 | 0.7 | <0.001 |
| Diseases of the respiratory system | 0.94 | 0.07 | 0.82 | 1.09 | 0.43 |
| Diseases of the skin and subcutaneous tissue | 0.28 | 0.22 | 0.18 | 0.44 | <0.001 |
| Endocrine, nutritional and metabolic diseases | 0.17 | 0.21 | 0.12 | 0.26 | <0.001 |
| Injury, poisoning and certain other consequences of external causes | 0.36 | 0.1 | 0.29 | 0.43 | <0.001 |
| Mental, Behavioral and Neurodevelopmental disorders | 0.45 | 0.47 | 0.18 | 1.14 | 0.09 |
| Neoplasms | 2.8 | 0.08 | 2.39 | 3.28 | <0.001 |
| Symptoms, signs and abnormal clinical and laboratory findings, not elsewhere classified | 0.1 | 0.14 | 0.07 | 0.13 | <0.001 |

**Table 5:** Multivariable logistic regression predicting emergency readmission (n = 52125). Results are presented as adjusted odds ratios, with 95% CI based on standard errors clustered at the patient level.

| **variable** | **estimate** | **SE** | **lower 95% CI** | **upper 95% CI** | **p-value** |
|---|---|---|---|---|---|
| (Intercept) | 0.06 | 0.12 | 0.05 | 0.08 | <0.001 |
| Atypical transfers | 1.00 | 0.03 | 0.95 | 1.05 | 0.92 |
| Regular transfers | 1.02 | 0.01 | 1.00 | 1.04 | 0.02 |
| Site transfers | 1.03 | 0.03 | 0.97 | 1.09 | 0.38 |
| *Reference aged 18 to 40 years* | | | | | |
| 40-65 | 1.19 | 0.04 | 1.09 | 1.3 | <0.001 |
| Over 65 | 1.17 | 0.04 | 1.07 | 1.28 | <0.001 |
| *Reference 0 Elixhauser comorbidities* | | | | | |
| Over 5 | 2.79 | 0.06 | 2.5 | 3.11 | <0.001 |
| 1-4 | 1.78 | 0.05 | 1.61 | 1.96 | <0.001 |
| *Reference not weekend admission* | | | | | |
| Weekend admission | 1.01 | 0.03 | 0.96 | 1.07 | 0.63 |

| | | | | | |
|---|---|---|---|---|---|
| *Reference Discharge not to usual place of residence* | | | | | |
| Usual place of residence | 2.15 | 0.05 | 1.95 | 2.37 | <0.001 |
| *Reference no ICU attendance* | | | | | |
| ICU admission | 0.74 | 0.06 | 0.66 | 0.83 | <0.001 |
| *Reference female* | | | | | |
| Male | 1.08 | 0.02 | 1.03 | 1.13 | <0.001 |
| *Reference 1 primary diagnosis* | | | | | |
| 2-3 | 1.08 | 0.04 | 1.00 | 1.15 | 0.04 |
| Over 3 | 0.92 | 0.24 | 0.58 | 1.46 | 0.73 |
| *Reference 0 imaging surgical procedures* | | | | | |
| 1-3 | 0.8 | 0.03 | 0.76 | 0.85 | <0.001 |
| 4-6 | 0.72 | 0.07 | 0.62 | 0.83 | <0.001 |
| Over 6 | 0.72 | 0.18 | 0.51 | 1.02 | 0.06 |
| *Reference 0 minor surgical procedures* | | | | | |
| 1-2 | 0.91 | 0.04 | 0.85 | 0.97 | 0.01 |
| 3-5 | 0.82 | 0.13 | 0.64 | 1.05 | 0.12 |
| Over 5 | 0.76 | 0.36 | 0.37 | 1.54 | 0.44 |
| *Reference 0 intermediate surgical procedures* | | | | | |
| 1-4 | 0.92 | 0.03 | 0.87 | 0.98 | 0.01 |
| 5-7 | 0.97 | 0.15 | 0.72 | 1.3 | 0.84 |
| 8-10 | 1.1 | 0.2 | 0.75 | 1.63 | 0.62 |
| Over 10 | 1.86 | 0.27 | 1.1 | 3.13 | 0.02 |
| *Reference 0 major surgical procedures* | | | | | |
| 1-3 | 0.77 | 0.04 | 0.71 | 0.83 | <0.001 |
| 4-6 | 0.74 | 0.16 | 0.54 | 1.02 | 0.07 |
| Over 6 | 0.89 | 0.37 | 0.43 | 1.86 | 0.76 |
| *Reference admission between 7am and 7pm* | | | | | |
| Out-of-hours admission | 0.94 | 0.02 | 0.9 | 0.99 | 0.01 |
| *Reference African ethnicity code* | | | | | |
| Any other Asian background | 0.93 | 0.08 | 0.79 | 1.09 | 0.36 |
| Any other Black background | 0.74 | 0.1 | 0.6 | 0.9 | <0.001 |
| Any other ethnic group | 0.83 | 0.07 | 0.73 | 0.94 | <0.001 |
| Any other mixed background | 1.03 | 0.16 | 0.76 | 1.39 | 0.87 |
| Any other White background | 0.86 | 0.07 | 0.75 | 0.98 | 0.02 |
| Bangladeshi | 0.91 | 0.16 | 0.66 | 1.25 | 0.55 |
| British | 1.04 | 0.06 | 0.92 | 1.17 | 0.56 |
| Caribbean | 1.02 | 0.07 | 0.88 | 1.18 | 0.79 |
| Chinese | 0.77 | 0.19 | 0.53 | 1.12 | 0.18 |
| Indian | 0.89 | 0.08 | 0.76 | 1.04 | 0.14 |
| Irish | 1.07 | 0.08 | 0.92 | 1.26 | 0.37 |
| Pakistani | 0.98 | 0.13 | 0.76 | 1.26 | 0.89 |
| Unknown | 0.71 | 0.07 | 0.63 | 0.82 | 0 |
| White and Asian | 0.53 | 0.29 | 0.3 | 0.93 | 0.03 |
| White and Black African | 1.06 | 0.27 | 0.63 | 1.79 | 0.82 |
| White and Black Caribbean | 1.56 | 0.16 | 1.15 | 2.12 | <0.001 |
| *Reference certain infectious and parasitic diseases ICD-10 chapter* | | | | | |
| Congenital malformations, deformations and chromosomal abnormalities | 0.45 | 0.74 | 0.11 | 1.93 | 0.28 |

| | | | | | |
|---|---|---|---|---|---|
| Diseases of the blood and blood-forming organs and certain disorders involving the immune mechanism | 2.33 | 0.08 | 1.98 | 2.72 | <0.001 |
| Diseases of the circulatory system | 0.67 | 0.05 | 0.6 | 0.74 | 0 |
| Diseases of the digestive system | 1.14 | 0.06 | 1.01 | 1.28 | 0.03 |
| Diseases of the ear and mastoid process | 0.78 | 0.35 | 0.4 | 1.55 | 0.48 |
| Diseases of the eye and adnexa | 0.61 | 0.27 | 0.36 | 1.04 | 0.07 |
| Diseases of the genitourinary system | 1.08 | 0.06 | 0.96 | 1.21 | 0.2 |
| Diseases of the musculoskeletal system and connective tissue | 0.95 | 0.09 | 0.8 | 1.13 | 0.58 |
| Diseases of the nervous system | 0.68 | 0.1 | 0.57 | 0.83 | <0.001 |
| Diseases of the respiratory system | 0.98 | 0.05 | 0.89 | 1.09 | 0.74 |
| Diseases of the skin and subcutaneous tissue | 0.92 | 0.09 | 0.77 | 1.1 | 0.37 |
| Endocrine, nutritional and metabolic diseases | 0.99 | 0.08 | 0.84 | 1.15 | 0.86 |
| Factors influencing health status and contact with health services | 0.81 | 0.24 | 0.5 | 1.3 | 0.38 |
| Injury, poisoning and certain other consequences of external causes | 0.73 | 0.06 | 0.65 | 0.82 | <0.001 |
| Mental, Behavioral and Neurodevelopmental disorders | 0.62 | 0.27 | 0.37 | 1.05 | 0.07 |
| Neoplasms | 1.65 | 0.06 | 1.46 | 1.87 | <0.001 |
| Symptoms, signs and abnormal clinical and laboratory findings, not elsewhere classified | 0.97 | 0.06 | 0.87 | 1.08 | 0.58 |
| *Reference LOS 0-1 days* | | | | | |
| 1-2 | 0.81 | 0.07 | 0.7 | 0.93 | <0.001 |
| 2-4 | 0.87 | 0.07 | 0.76 | 0.99 | 0.04 |
| 4-6 | 0.91 | 0.07 | 0.8 | 1.05 | 0.2 |
| 6-8 | 1.02 | 0.07 | 0.89 | 1.17 | 0.79 |
| 8-10 | 1.15 | 0.07 | 0.99 | 1.33 | 0.06 |
| Over 10 | 1.31 | 0.07 | 1.15 | 1.49 | <0.001 |

**Sensitivity analyses:**

**Length of stay**

Table 6a: Adjusted average marginal effects for the relationship between atypical transfers, regular transfers, site transfers and increased length of stay derived by a gamma GLM with log link, from observations whose standardised residuals were below 3 (n = 55003)

| **Variable** | **AME** | **SE** | **Lower 95%CI** | **Upper 95%CI** | **p-value** |
|---|---|---|---|---|---|
| Atypical transfers | 2.69 | 0.13 | 2.44 | 2.44 | <0.001 |
| Regular transfers | 1.93 | 0.05 | 1.83 | 2.03 | <0.001 |
| Site transfers | 3.01 | 0.15 | 2.73 | 3.30 | <0.001 |

Table 6b: Adjusted average marginal effects for the relationship between atypical transfers, regular transfers, site transfers and increased length of stay derived by a gamma GLM with log link, from spells in which the patient was discharged alive and did not self-discharge (n=51210)

| Variable | AME | SE | Lower 95%CI | Upper 95%CI | p-value |
|---|---|---|---|---|---|
| Atypical transfers | 2.80 | 0.15 | 2.51 | 3.09 | <0.001 |
| Regular transfers | 1.88 | 0.06 | 1.77 | 1.99 | <0.001 |
| Site transfers | 2.93 | 0.17 | 2.60 | 3.26 | <0.001 |

Table 6c: Adjusted average marginal effects for the relationship between atypical transfers, regular transfers, site transfers and increased length of stay derived by a gamma GLM with log link, with BUPA surgical categorisations used for procedure adjustment (n=55152).

| Variable | AME | SE | Lower 95%CI | Upper 95%CI | p-value |
|---|---|---|---|---|---|
| Atypical transfers | 3.03 | 0.15 | 2.75 | 3.32 | <0.001 |
| Regular transfers | 2.13 | 0.05 | 2.03 | 2.24 | <0.001 |
| Site transfers | 3.25 | 0.17 | 2.91 | 3.59 | <0.001 |

Table 6d: Adjusted average marginal effects for the relationship between atypical transfers, regular transfers, site transfers and increased length of stay derived by a gamma GLM with log link, from spells in which the patient was free of infection or colonisation for the duration of their spell (n=47708)

| Variable | AME | SE | Lower 95%CI | Upper 95%CI | p-value |
|---|---|---|---|---|---|
| Atypical transfers | 2.91 | 0.16 | 2.60 | 3.21 | <0.001 |
| Regular transfers | 1.95 | 0.06 | 1.83 | 2.07 | <0.001 |
| Site transfers | 3.07 | 0.18 | 2.72 | 3.42 | <0.001 |

Table 6e: Adjusted average marginal effects for the relationship between atypical transfers, regular transfers, site transfers and increased length of stay derived by a gamma GLM with log link, with approximate month of admission included in the adjustment set (n=55152)

| Variable | AME | SE | Lower 95%CI | Upper 95%CI | p-value |
|---|---|---|---|---|---|
| Atypical transfers | 2.81 | 0.14 | 2.52 | 3.10 | <0.001 |
| Regular transfers | 1.93 | 0.05 | 1.82 | 2.04 | <0.001 |
| Site transfers | 3.02 | 0.17 | 2.70 | 3.35 | <0.001 |

Table 6f: Adjusted average marginal effects for the relationship between atypical transfers, regular transfers, site transfers and increased length of stay derived by a gamma GLM with log link, with alternative ward speciality categorisations obtained by using the *first* TFC associated with spells (n=55152)

| Variable | AME | SE | Lower 95%CI | Upper 95%CI | p-value |
|---|---|---|---|---|---|
| Atypical transfers | 2.58 | 0.15 | 2.28 | 2.87 | <0.001 |
| Regular transfers | 1.97 | 0.05 | 1.86 | 2.07 | <0.001 |
| Site transfers | 3.03 | 0.17 | 2.7 | 3.35 | <0.001 |

**Mortality**

Table 7a: Adjusted odds ratios for the relationship between atypical transfers, regular transfers, site transfers and mortality derived by a logistic regression, from observations whose standardised residuals were below 3 (n = 54981)

| **Variable** | **Variable** | **SE** | **Lower 95%CI** | **Upper 95%CI** | **p-value** |
|---|---|---|---|---|---|
| Atypical transfers | 0.96 | 0.04 | 0.89 | 1.04 | 0.38 |
| Regular transfers | 0.98 | 0.01 | 0.96 | 1.01 | 0.23 |
| Site transfers | 0.71 | 0.06 | 0.63 | 0.80 | <0.001 |

Table 7b: Adjusted odds ratios for the relationship between atypical transfers, regular transfers, site transfers and mortality derived by a logistic regression, with BUPA surgical categorisations used for procedure adjustment (n=55152).

Bupa surgical code categorisation:

| **Variable** | **Variable** | **SE** | **Lower 95%CI** | **Upper 95%CI** | **p-value** |
|---|---|---|---|---|---|
| Atypical transfers | 0.98 | 0.04 | 0.91 | 1.06 | 0.67 |
| Regular transfers | 0.99 | 0.01 | 0.97 | 1.02 | 0.61 |
| Site transfers | 0.73 | 0.06 | 0.65 | 0.82 | <0.001 |

Table 7c: Adjusted odds ratios for the relationship between atypical transfers, regular transfers, site transfers and mortality derived by a logistic regression, from spells in which the patient was free of infection or colonisation for the duration of their spell (n=47584)

| **Variable** | **Variable** | **SE** | **Lower 95%CI** | **Upper 95%CI** | **p-value** |
|---|---|---|---|---|---|
| Atypical transfers | 0.96 | 0.04 | 0.88 | 1.04 | 0.29 |
| Regular transfers | 0.99 | 0.01 | 0.96 | 1.02 | 0.47 |
| Site transfers | 0.74 | 0.06 | 0.66 | 0.84 | <0.001 |

Table 7d: Adjusted odds ratios for the relationship between atypical transfers, regular transfers, site transfers and mortality derived by a logistic regression, with approximate month of admission included in the adjustment set (n=55152)

| **Variable** | **Variable** | **SE** | **Lower 95%CI** | **Upper 95%CI** | **p-value** |
|---|---|---|---|---|---|
| Atypical transfers | 0.96 | 0.04 | 0.88 | 1.04 | 0.29 |
| Regular transfers | 0.99 | 0.01 | 0.96 | 1.02 | 0.41 |
| Site transfers | 0.71 | 0.06 | 0.63 | 0.80 | <0.001 |

Table 7e: Adjusted odds ratios for the relationship between atypical transfers, regular transfers, site transfers and mortality derived by a logistic regression, with alternative ward speciality categorisations obtained by using the *first* TFC associated with spells (n= 55000)

| **Variable** | **Variable** | **SE** | **Lower 95%CI** | **Upper 95%CI** | **p-value** |
|---|---|---|---|---|---|
| Atypical transfers | 0.90 | 0.05 | 0.81 | 0.99 | 0.03 |
| Regular transfers | 0.99 | 0.01 | 0.97 | 1.02 | 0.52 |
| Site transfers | 0.72 | 0.06 | 0.64 | 0.80 | <0.001 |

**Readmission**

The 30-day emergency readmission model did not obtain any observations whose standardised residuals were below 3, therefore this sensitivity analysis was omitted.

Table 8a: Adjusted odds ratios for the relationship between atypical transfers, regular transfers, site transfers and 30-day emergency readmission, with BUPA surgical categorisations used for procedure adjustment (n=52125).

| Variable | Estimate | SE | Lower 95%CI | Upper 95%CI | p-value |
|---|---|---|---|---|---|
| Atypical transfers | 1.00 | 0.03 | 0.95 | 1.06 | 0.89 |
| Regular transfers | 1.01 | 0.01 | 0.99 | 1.03 | 0.22 |
| Site transfers | 1.02 | 0.03 | 0.96 | 1.08 | 0.46 |

Table 8b: Adjusted odds ratios for the relationship between atypical transfers, regular transfers, site transfers and 30-day emergency readmission, from spells in which the patient was free of infection or colonisation for the duration of their spell (n=44988)

| Variable | Estimate | SE | Lower 95%CI | Upper 95%CI | p-value |
|---|---|---|---|---|---|
| Atypical transfers | 1.00 | 0.03 | 0.94 | 1.06 | 0.99 |
| Regular transfers | 1.03 | 0.01 | 1.01 | 1.05 | 0.01 |
| Site transfers | 1.00 | 0.04 | 0.93 | 1.07 | 0.96 |

Table 8c: Adjusted odds ratios for the relationship between atypical transfers, regular transfers, site transfers and 30-day emergency readmission, with approximate month of admission included in the adjustment set (n=52125)

| Variable | Estimate | SE | Lower 95%CI | Upper 95%CI | p-value |
|---|---|---|---|---|---|
| Atypical transfers | 1.00 | 0.03 | 0.95 | 1.05 | 0.90 |
| Regular transfers | 1.02 | 0.01 | 1.00 | 1.04 | 0.02 |
| Site transfers | 1.03 | 0.03 | 0.97 | 1.09 | 0.37 |

Table 8d: Adjusted odds ratios for the relationship between atypical transfers, regular transfers, site transfers and 30-day emergency readmission, with alternative ward speciality categorisations obtained by using the *first* TFC associated with spells (n= 52125).

| Variable | Estimate | SE | Lower 95%CI | Upper 95%CI | p-value |
|---|---|---|---|---|---|
| Atypical transfers | 1.00 | 0.03 | 0.95 | 1.05 | 0.92 |
| Regular transfers | 1.02 | 0.01 | 1.00 | 1.04 | 0.02 |
| Site transfers | 1.03 | 0.03 | 0.97 | 1.09 | 0.38 |

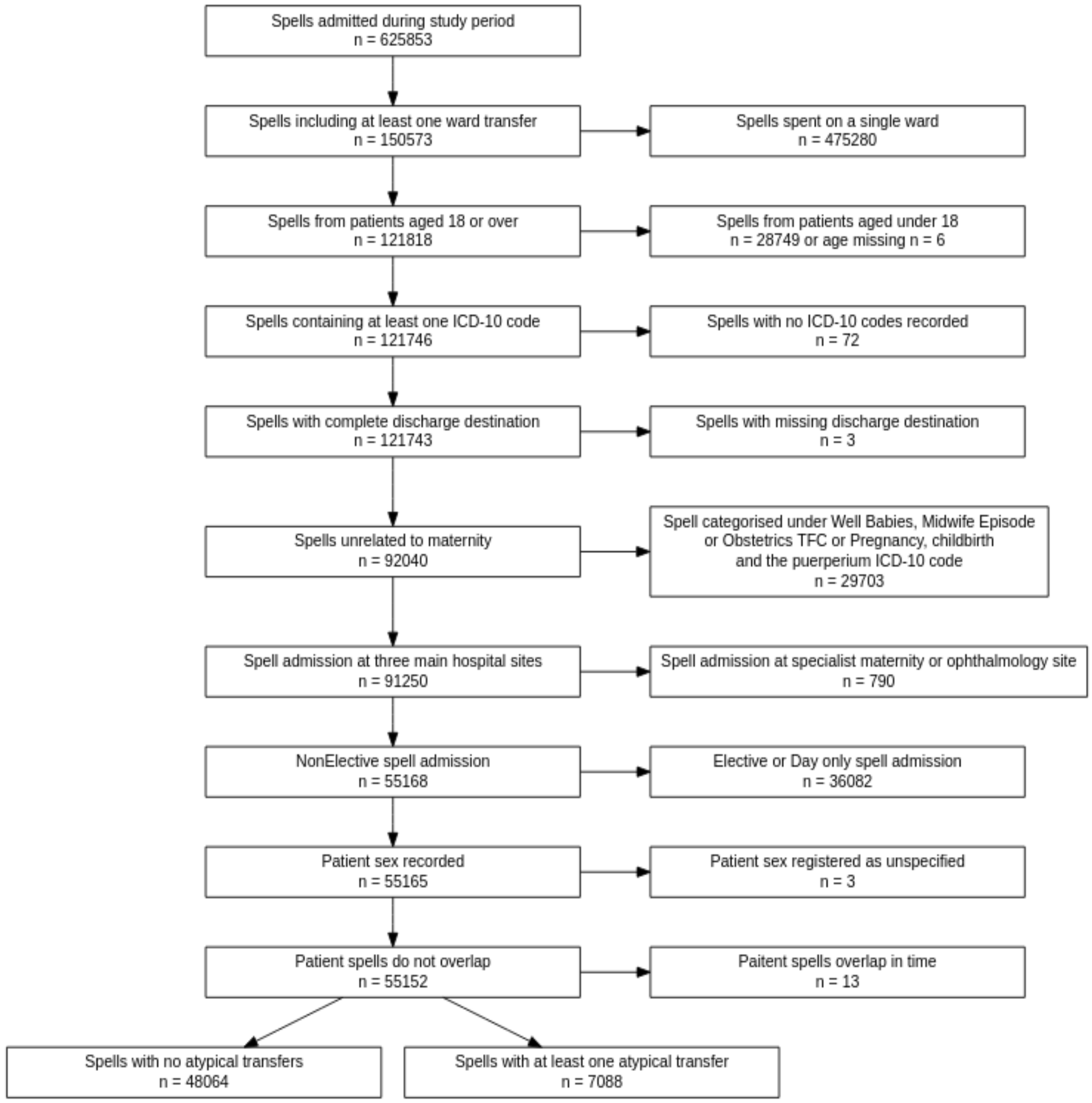


**Figure 1:** A flow diagram summarising the inclusion and exclusion criteria for the study participants. The final sample included 55,152 patient spells pertaining to 38,101 individual patients. Of these 7,088 (12.9%) spells included at least one atypical transfer.

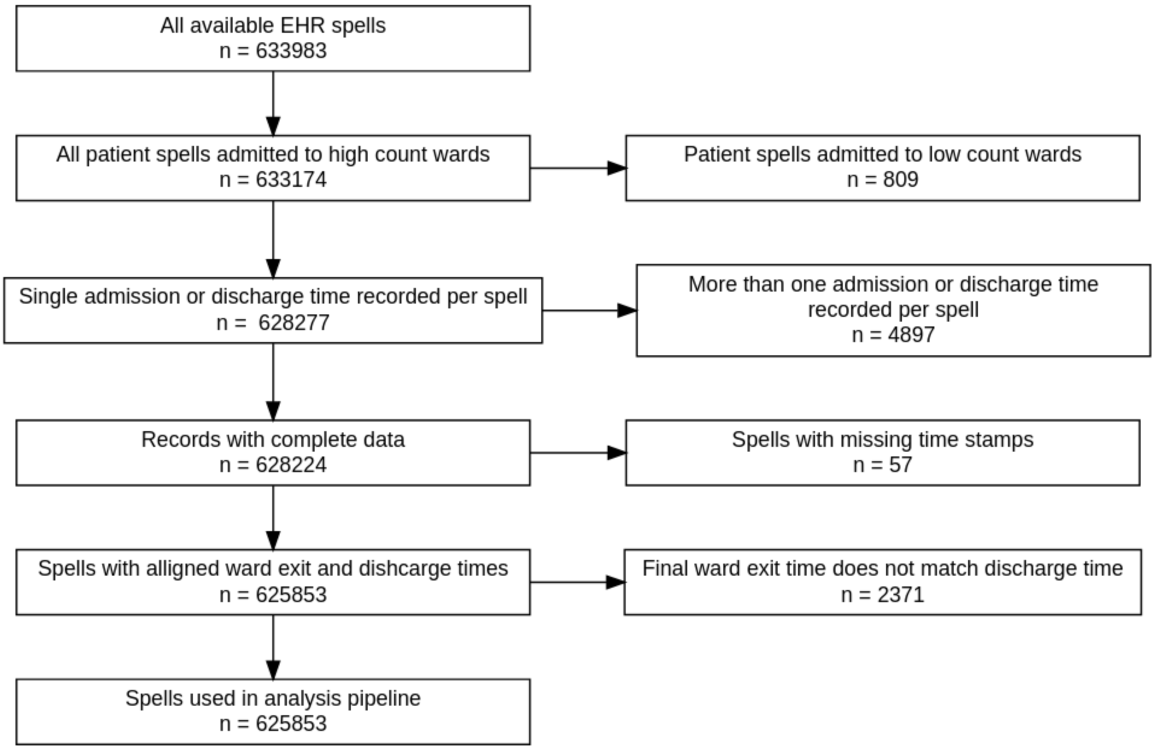


Figure 2: A flow diagram showing data cleaning decisions and the number of spells they affected prior to analysis.

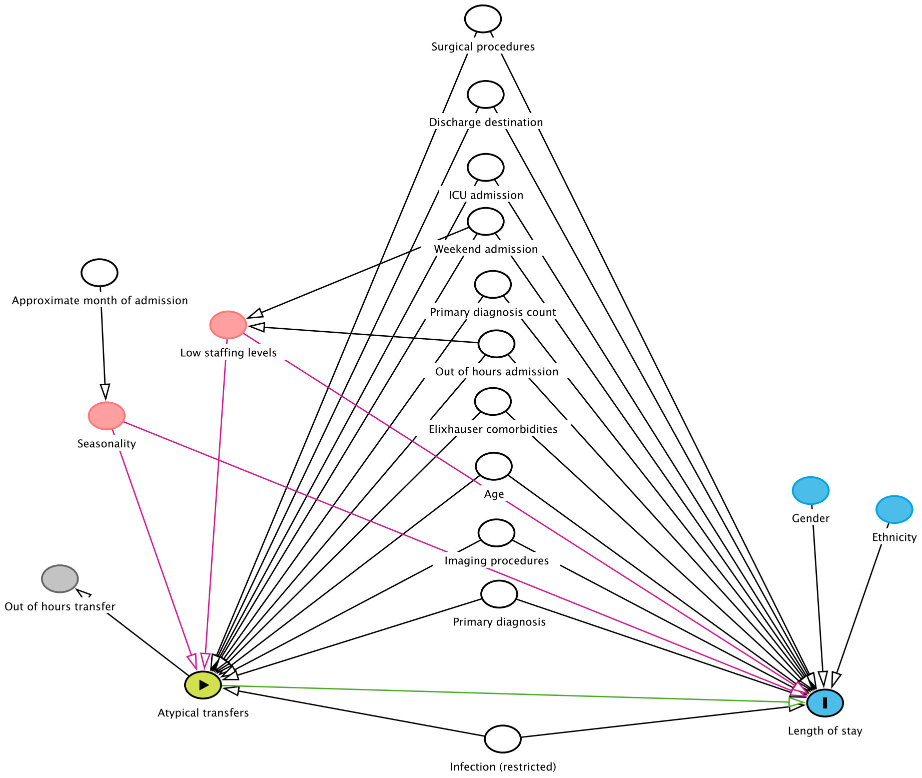


Figure 3: Example of a Directed Acyclic Graph (DAG) exploring the relationships between atypical transfers and length of stay.

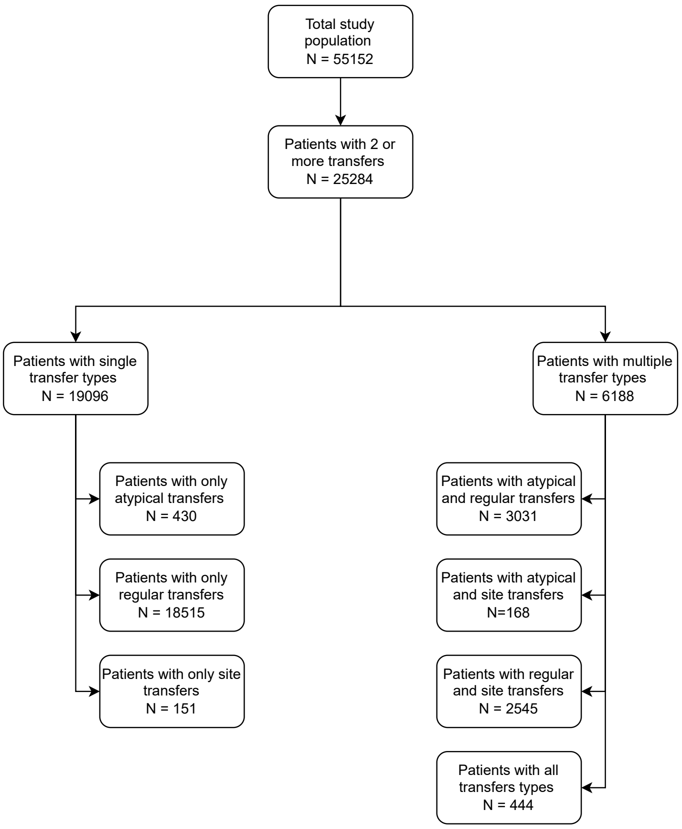


Figure 4: A breakdown of transfer type for patients with multiple transfers.

**Note 1a: Identifying pregnant women**

Women attending the hospital for reasons related to pregnancy were removed from the analysis if incurring the final TFC code *well babies*, *midwife episode* or *obstetrics* or the first chapter code *Pregnancy, childbirth and the puerperium.*

**Note 1b: Categorisation of surgical procedures**
Abbott et al., (2011) defined code lists of 'inclusive' procedures that could be considered surgery, such as minor surgery, interventional radiology procedures and diagnostic endoscopies.[2] The second 'intermediate' category included more severe procedures routinely undertaken in an operating theatre and/or under general or regional anaesthesia, while third 'restrictive' category included major procedures, defined as a complex procedure which may result in in tissue injury. While the authors originally defined categories not mutually exclusive (e.g. a restrictive procedure would also be counted in the inclusive category), in our analysis the codes were separated such that inclusive, intermediate and restrictive categories were distinct.

**Note 1c: Alternative categorisation of surgical procedures by Bupa Schedule of Procedures**

The Clinical Coding and Schedule Development Group (CCSD) schedule was developed to allow clinicians in private practice to be reimbursed for their work in the private healthcare sector. Surgical codes are allocated a complexity level (minor, intermediate, major, major plus and complex major) with an associated cost. While OPCS and CCSD codes cover the same major areas (or chapters), mapping OPCS and CCSD codes is challenging and cannot be achieved through a direct match. OPCS has more than 6,000 codes while CCSD has around 2,070, and therefore many OPCS codes may be associated with one CCSD code.

The Bupa Schedule of Procedures updated on 15 Oct 2021 by BUPA healthcare was used as a source of CCSD categories.[1] The 'hospital category' classification was selected to allocate procedures a severity level. The codes were matched to OPCS-4 codes through an exact match on the first 2 figures. A fuzzy match allowing a distance of 1 was used to match the remaining code figures to OPCS 4 codes. This resulted in a match for 2056 codes of the 5941 unique OPCS seen in the dataset.
